# Estimating the transmissibility of SARS-CoV-2 during periods of high, low and zero case incidence

**DOI:** 10.1101/2021.11.28.21264509

**Authors:** Nick Golding, David J. Price, Gerard E. Ryan, Jodie McVernon, James M. McCaw, Freya M. Shearer

## Abstract

Against a backdrop of widespread global transmission, a number of countries have successfully brought large outbreaks of COVID-19 under control and maintained near-elimination status. A key element of epidemic response is the tracking of disease transmissibility in near real-time. During major outbreaks, the reproduction rate can be estimated from a time-series of case, hospitalisation or death counts. In low or zero incidence settings, knowing the potential for the virus to spread is a response priority. Absence of case data means that this potential cannot be estimated directly.

We present a semi-mechanistic modelling framework that draws on time-series of both behavioural data and case data (when disease activity is present) to estimate the transmissibility of SARS-CoV-2 from periods of high to low – or zero – case incidence, with a coherent transition in interpretation across the changing epidemiological situations. Of note, during periods of epidemic activity, our analysis recovers the effective reproduction number, while during periods of low – or zero – case incidence, it provides an estimate of transmission risk. This enables tracking and planning of progress towards the control of large outbreaks, maintenance of virus suppression, and monitoring the risk posed by re-introduction of the virus.

We demonstrate the value of our methods by reporting on their use throughout 2020 in Australia, where they have become a central component of the national COVID-19 response.

## Introduction

The first 12 months of the COVID-19 pandemic led to overwhelmed health systems and enormous social disruption across the globe. Government strategy and public responses to COVID-19 have been highly variable. Prior to the global circulation of the Delta variant, a small number of jurisdictions had achieved extended periods of elimination through 2020 and into early 2021, including Taiwan, Thailand, New Zealand and Australia [1, 2, 3]. Meanwhile, parts of Europe and the Americas were heavily impacted by COVID-19 [4, 5], with health systems overwhelmed by multiple explosive outbreaks. The Delta variant — with its increased transmissibility — has recently led to epidemic activity, now likely to be sustained, in a number of previously low prevalence settings [6, 7].

A key element of epidemic response is the close monitoring of the rate of disease spread, via estimation of the effective reproduction number (*R*_eff_) — the average number of new infections caused by an infected individual in the presence of public health interventions and where no assumption of 100% susceptibility is made. Methods are well-established for near real-time estimation of this critical value and estimates are routinely assessed by decision-makers through the course of an epidemic [8, 9, 10, 11, 12]. When *R*_eff_ is above 1, the epidemic is estimated to be growing. If control measures, population immunity, or other factors can bring *R*_eff_ below 1, then the epidemic is estimated to be in decline. Accurate and timely estimation of *R*_eff_, and the timely adjustment of interventions in response to it, is critical for the sustainable and successful management of COVID-19.

However, when incident cases are driven to very low levels — as occurred in Australia following the first wave of COVID-19 from February to April 2020 — established methods for estimating *R*_eff_ are no longer informative. Yet the virus remains a threat, as evidenced by multiple instances of re-introduction and subsequent additional waves in Australia throughout 2020 and early 2021. Independent of whether local (and temporary) elimination is achieved, knowledge of SARS-CoV-2’s potential transmissibility and the risk of resurgence is a response priority.

Here, by making use of social and behavioural data, we demonstrate a novel method for estimating the ability of the virus to spread in a population, which is informative even when case incidence is very low or zero. In the absence of cases, our method estimates the ability of the virus, if it were present, to spread in a population, which we define as the ‘transmission potential’. We use the word ‘potential’ to distinguish this quantity from an estimate of actual transmission. When the virus is present, our method recovers the effective reproduction number and, additionally, the deviation between the *R*_eff_ and the transmission potential. Applying this method in real-time provides an estimate of the transmissibility of SARS-CoV-2 in periods of high, low, and even zero, case incidence, with a coherent and seamless transition in interpretation across the changing epidemiological situations.

Our innovative methods and workflows address a major challenge in epidemic situational awareness: assessing epidemic risk when case numbers are driven to low levels or (temporary) elimination is achieved, as frequently occurred in Australia through 2020–21 [3]. We have routinely applied this method to all Australian states and territories and reported the outputs to peak national decision-making committees on a weekly basis since early May 2020. The concepts of transmission potential and *R*_eff_ have been incorporated into key instruments of government, including Australia’s national COVID-19 surveillance plan [13]. The transmission potential and *R*_eff_ are reported to the public through the Australian Government’s weekly Common Operating Picture [14]. While not addressed in this article, our methods have recently been updated to include consideration of variants of concern [3] and the effect of vaccination [14] on reducing the ability of the virus to spread in the population.

### Novel method for estimating temporal trends in the transmissibility of SARS-CoV-2

The effective reproduction number is the product of the number of contacts an infectious person makes and the per contact probability of infection (the latter of which depends on the nature and duration of contact) [15]. Both quantities are impacted by changes in behaviour, which are in turn driven by changes in policy, such as stay-at-home orders and handwashing advice, and the population’s perception and evaluation of risk, among other factors. The new techniques introduced here provide an estimate for how observed changes in rates of social contact and the per contact probability of infection translate to changes in the ability of the virus to spread.

We estimate the time-varying ability of SARS-CoV-2 to spread in a population using a novel semi-mechanistic model informed by data on cases, population behaviours and health system effectiveness (see Materials and Methods). We separately model transmission from locally acquired cases (local-to-local transmission) and from overseas acquired cases (import-to-local transmission). We model local-to-local transmission (*R*_eff_) using two components (Figure 1): the average population-level trend in *R*_eff_ driven by interventions that primarily target transmission from local cases, specifically changes in physical distancing behaviour and case targeted measures (Component 1, the ‘transmission potential’ or TP); and short-term fluctuations in *R*_eff_ to capture stochastic dynamics of transmission, such as clusters of cases and short periods of lower-than-expected transmission (Component 2, the ‘deviation’ between TP and *R*_eff_). During periods of low or zero transmission, TP provides an evaluation of the ability of the virus to spread, informing risk-assessments and supporting public health planning and response [16].

**Figure 1:**
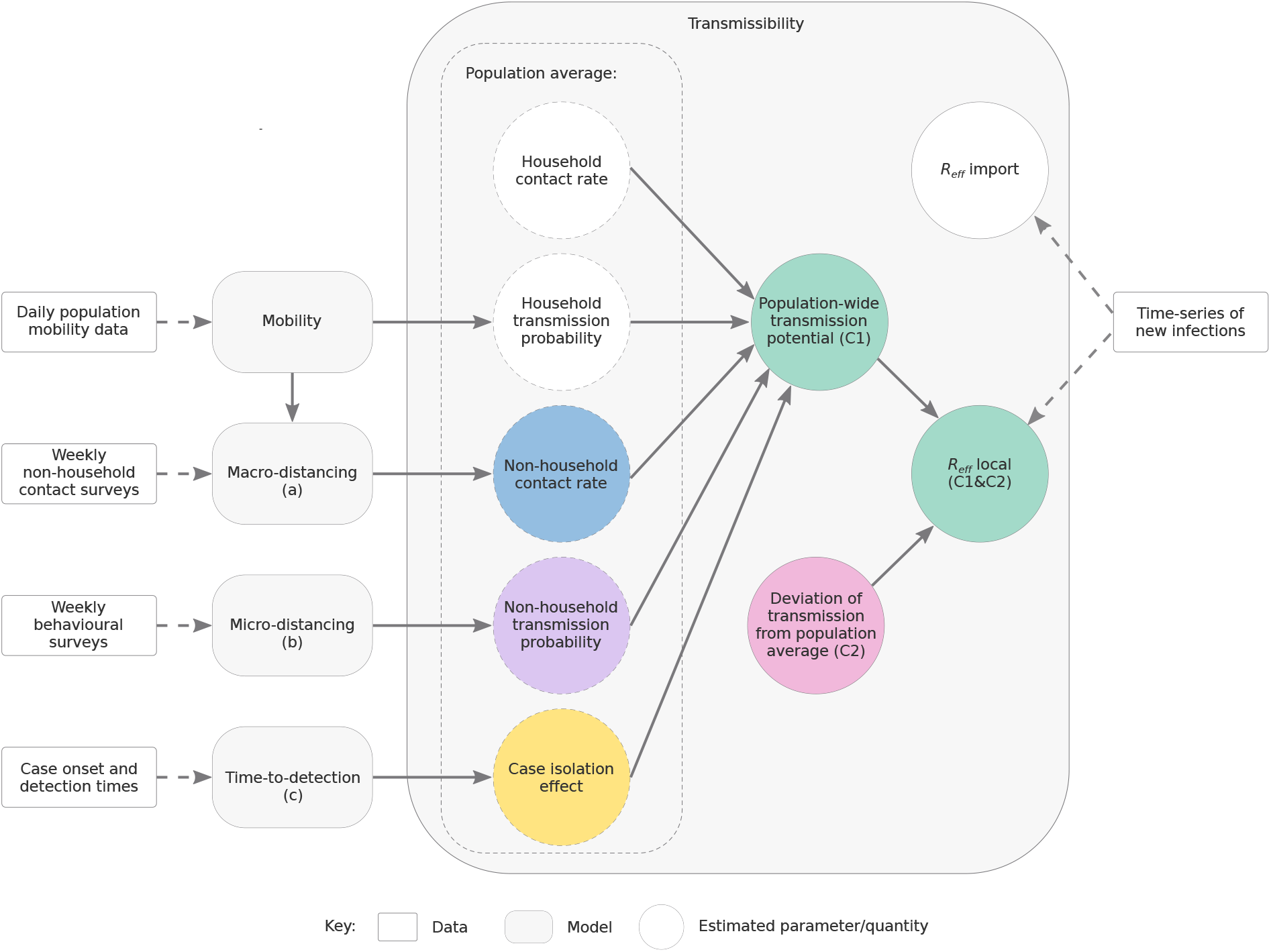
Depiction of the relationship between data sources, model components, and reported quantities.

To estimate Component 1, we use three sub-models (Figure 1, labelled a, b and c). We distinguish between two types of physical distancing behaviour:

a. macro-distancing, defined as the reduction in the average rate of non-household contacts, and assessed through weekly nationwide surveys of the daily number of non-household contacts; and
b. micro-distancing, defined as the reduction in transmission probability per non-household contact, and assessed through weekly nationwide surveys from which we estimate the proportion of the population reporting always keeping 1.5 metre physical distance from non-household contacts.

By synthesising data from these surveys and numerous population mobility data streams made available by technology company Google, we infer temporal trends in macro- and micro-distancing behaviour (sub-models a and b). Furthermore, using data on the number of days from symptom onset to case notification for cases, we estimate the proportion of cases that are detected (and thus advised to isolate) by each day post-infection. By quantifying the temporal change in the probability density for the time-to-detection (sub-model c), the model estimates how earlier isolation of cases — due to improvements in contact tracing, expanded access to testing, more inclusive case definitions, and other factors impacting detection rates — reduces the ability of SARS-CoV-2 to spread.

Transmission potential (Component 1) reflects the average potential for the virus to spread at the population level. During times of disease activity, Component 2 measures how transmission within the sub-populations that have the most active cases at a given point in time differs compared to that expected from the population-wide TP. The combination of Components 1 and 2 recovers the estimated *R*_eff_ (see Equation (10) in Materials and Methods), as per established methods [9, 10, 11]. When Component 2, the deviation between TP and *R*_eff_, is positively biased (*R*_eff_ *>* TP), it may indicate that transmission is concentrated in populations with higher-than-average levels of mixing, such as healthcare workers or meat processing workers. If negatively biased (*R*_eff_ *<* TP), it reflects suppressed transmission compared to expectation. This may be due to an effective public health response actively suppressing transmission (*e*.*g*., through test, trace, isolation and quarantine), or other factors such as local depletion of susceptible individuals, and/or the virus circulating in a sub-population with fewer-than-average social contacts.

### Assessing epidemic activity and risk in Australia

To demonstrate the utility of our method, we report on its application to Australian data on cases, population behaviour and health system effectiveness from the first 12 months of the COVID-19 pandemic. We focus on the period from early March 2020 to late January 2021 prior to emergence of variants of concern in Australia (first Alpha, then Delta) and vaccination roll out (refer to our recent technical report for details on our approach to variants of concern [3]). We describe our results in the context of the COVID-19 epidemiology and public health response in Australia during this period, noting that the methods were developed and applied during the pandemic and contributed to government response efforts. We report retrospective estimates (using data as of 24 January 2021 and our model as of September 2021). Where relevant, we also report estimates made at the time of analysis in 2020, which may differ as a result of updates to the case data and methodological improvements to our model over time, as well as minor statistical variation and smoothing.

Across its eight states and territories, Australia has managed a number of distinct phases of the pandemic — from an initial wave of importations (February–April 2020), to sustained periods of zero local case incidence (April–June 2020 and October–December 2020) to widespread community transmission (June–October 2020). Like elsewhere in the world, key interventions have included quarantine of overseas arrivals, restrictions on mobility and gathering sizes, advice on personal hygiene, and case targeted interventions. The specific measures, and the level of control of SARS-CoV-2 transmission, has varied between states and over time, according to changing epidemiology and response objectives, among other factors. The model has proven informative across vastly different and rapidly changing phases of the pandemic.

To highlight these different epidemiological situations and the insights gained from our model-based analysis, we draw on exemplar events from the Australian epidemic when describing our results below. In Table 1, we summarise the key types of information provided by estimated quantities under different epidemiological situations. Further, in Supplementary Figures S1–S7, we provide time-series estimates of each metric and model sub-component from early March 2020 to late January 2021 for each Australian state and territory.

**Table 1:** Definitions and interpretation of key estimated quantities from the model of SARS-CoV-2 transmissibility in different epidemiological contexts. *R*_eff_ = the effective reproduction number. TP = transmission potential. C2 = model component 2.

| Metric | Definition | Interpretation |  |
| --- | --- | --- | --- |
|  |  | Community transmission | Local elimination |
| TP | Expected reproduction number of a pathogen in the general population | If established in the general population, whether the epidemic is expected to grow ($TP > 1$) or decline ($TP < 1$) | Suitability for the pathogen, if it were present, to establish and maintain community transmission ($TP > 1$) or otherwise ($TP < 1$). |
| $R_{\text{eff}}$ | Average number of new infections caused by an infectious individual drawn from the active cases | Whether the epidemic is growing ($R_{\text{eff}} > 1$) or in decline ($R_{\text{eff}} < 1$) | Not applicable |
| C2 | Deviation between TP and $R_{\text{eff}}$ | Whether the virus is spreading faster (C2 positively biased) or slower (C2 negatively biased) among active cases than expected | Not applicable |

#### i) Initial wave of importations

Australia took an early and precautionary approach to managing the risk of importation of SARS-CoV-2. On 1 February 2020, when China was the only country reporting uncontained transmission, Australia restricted all travel from mainland China to Australia. Only Australian citizens and residents were permitted entry from mainland China. These individuals were advised to self-quarantine for 14 days from their date of arrival. From 20 March 2020, Australia closed its borders to all foreign nationals, and from 27 March, shifted to mandatory state-managed quarantine for returned citizens and residents, with weekly quotas on the number of arrivals. These policies remained in place at the time of writing.

During the first half of March 2020, *i*.*e*., prior to the border closure, daily case incidence increased sharply. Although more than two thirds of these cases had acquired their infection overseas, pockets of local transmission were reported in Australia’s largest cities of Sydney (New South Wales) and Melbourne (Victoria) [17] (Figure 2, panels A and E). From 16 March 2020, state governments progressively implemented — in rapid succession — a range of physical distancing measures to reduce and prevent community transmission. These measures were part of a coordinated national response strategy. By 31 March, Australians were strongly advised to leave their homes only for limited essential activities and public gatherings were limited to two people (known as “stay-at-home” restrictions). Health authorities also advised individuals to keep 1.5 metres distance from non-household members from mid-March [18].

**Figure 2:**
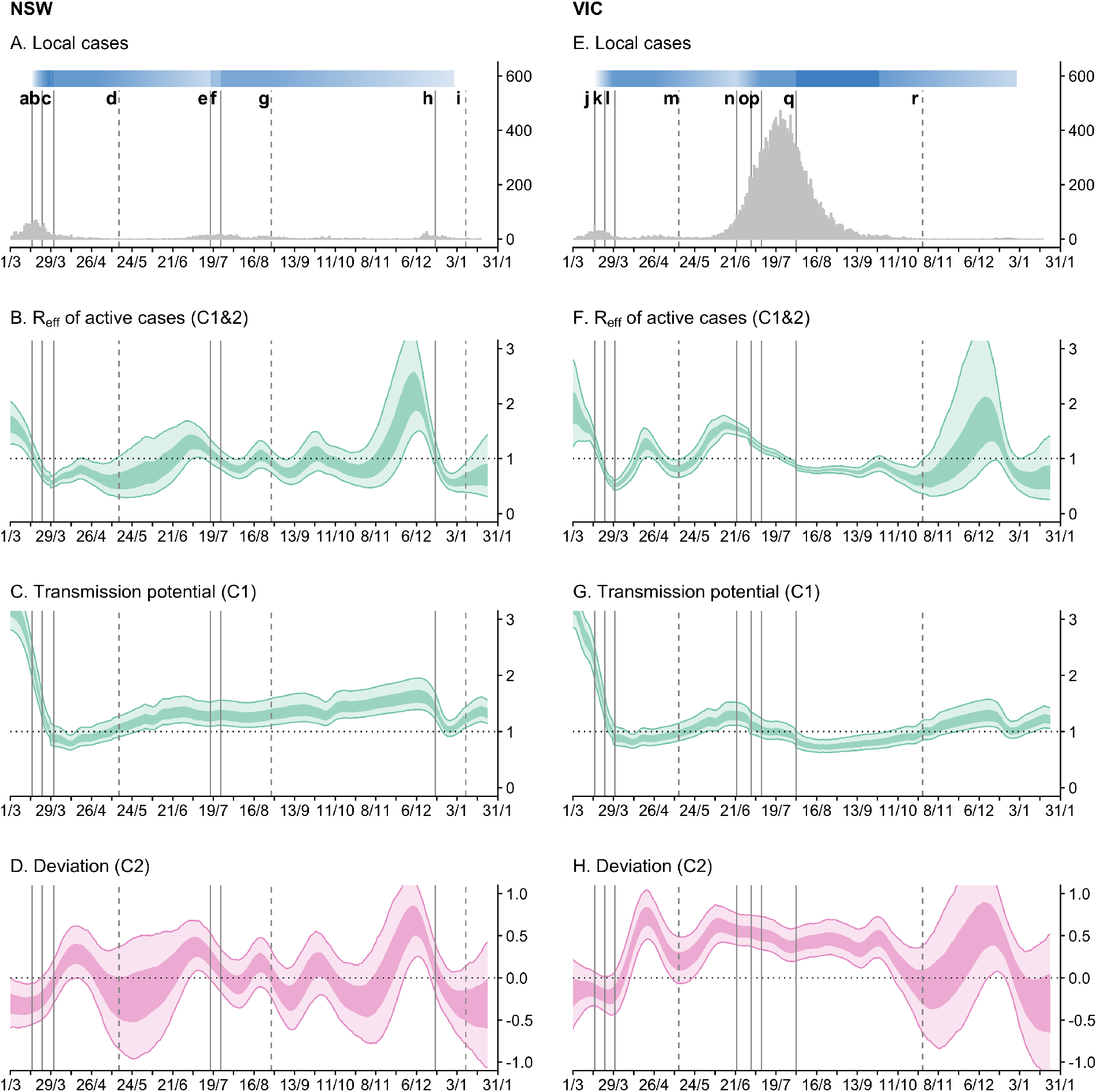
Time-series of daily local cases and transmissibility model components for the states of New South Wales (NSW) and Victoria (VIC) from 1 March 2020 to 24 January 2021. Light ribbons = 90% credible intervals; dark ribbons = 50% credible intervals. Vertical lines represent dates of key changes in restrictions on gatherings and movement, detailed in Table S1 (solid lines = tightening of restrictions; dashed lines = easing of restrictions). The blue bar is shaded according to the level of restrictions (lighter blue = less restrictions; darker blue = more restrictions). **A and E:** Daily new local cases by inferred infection date. **B and F:** State-wide local transmission potential (Component 1). **C and G:** *R*_eff_ of local active cases (Component 1&2). **D and H:** Deviation between transmission potential and *R*_eff_ (Component 2).

Through the second half of March 2020, we estimate that transmission potential across states and territories decreased substantially and rapidly from well above 1 to just below 1 (Figure 2, panels B and F). This reflected a marked increase in macro-/micro-distancing behaviour (Figure 3, panels B, C, F and G) and a decrease in time-to-case-detection (Figure 3, panels D and H). Our method, with its ability to distinguish between import-to-local and local-to-local transmission, estimates that the local *R*_eff_ dropped below 1 on 20 March in Victoria and 19 March in New South Wales — prior to the activation of stay-at-home restrictions (Figure 2, panels C and G). Physical distancing measures were implemented proactively — prior to the establishment of widespread community transmission — suggesting that the effect of these measures, in combination with border measures and case-targeted interventions, led to the definitive control of a first epidemic wave.

**Figure 3:**
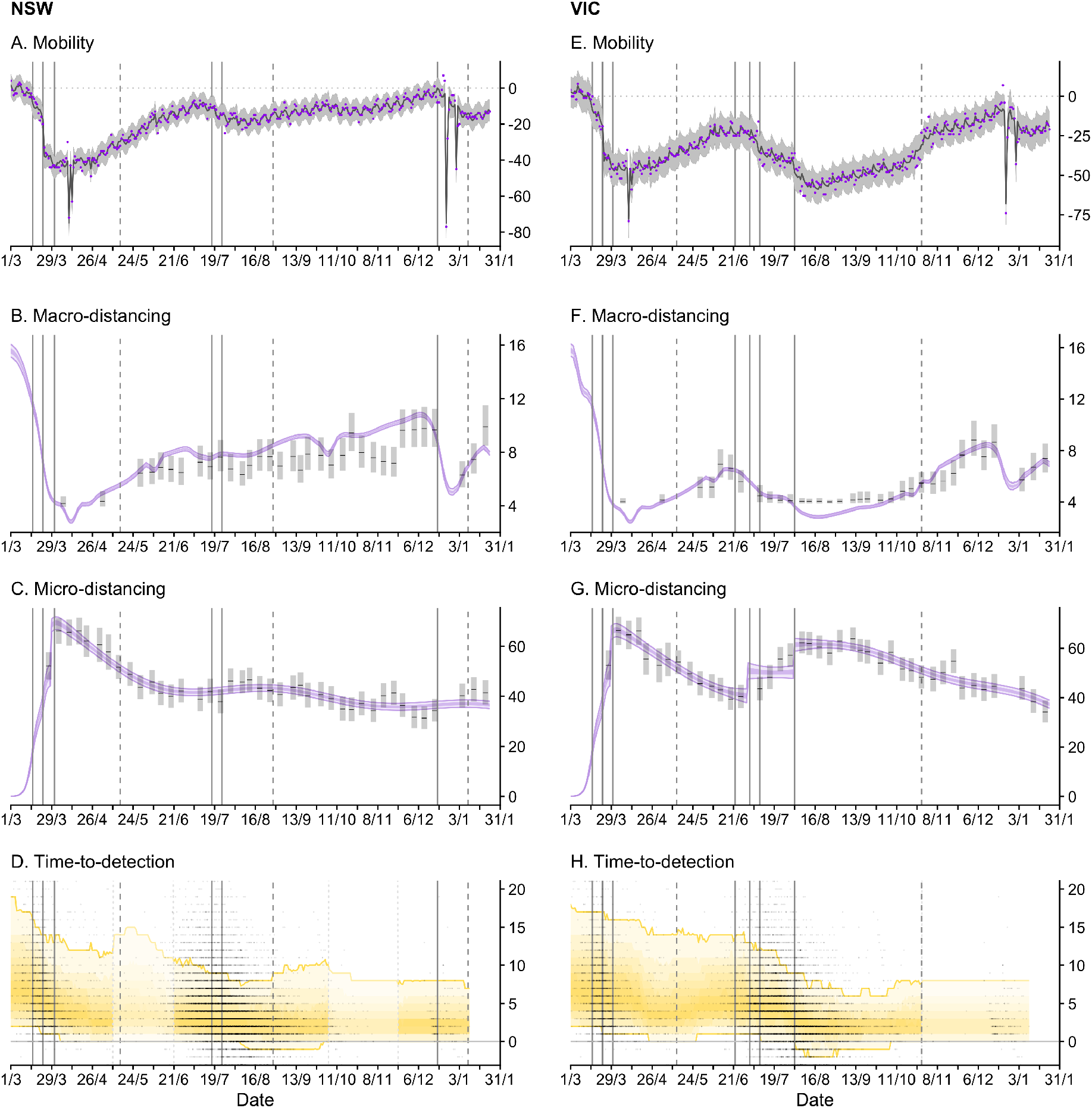
Time-series of each sub-model of transmission potential (Component 1) for New South Wales (NSW) and Victoria (VIC) from 1 March 2020 to 24 January 2021. Vertical lines represent dates of key changes in restrictions on gatherings and movement, detailed in Table S1 (solid lines = tightening of restrictions; dashed lines = easing of restrictions). The blue bar is shaded according to the level of restrictions (lighter blue = less restrictions; darker blue = more restrictions). **A and E:** Percentage change compared to a pre-COVID-19 baseline of one key population mobility data stream ‘Google: time a retail and recreation’. Purple dots are data stream values (percentage change on baseline). Solid lines and grey shaded regions are the estimated trend and 95% error interval estimated by our model. **B and F:** Estimated trends in macro-distancing behaviour, *i*.*e*., reduction in the daily rate of non-household contacts (dark purple ribbons = 50% credible intervals; light purple ribbons = 90% credible intervals). Estimates are informed by state-level data from nationwide weekly surveys (indicated by the black lines and grey rectangles) and population mobility data. **C and G:** Estimated trends in micro-distancing behaviour, *i*.*e*., reduction in transmission probability per non-household contact (dark purple ribbons = 50% credible intervals, light purple ribbons = 90% credible intervals). Estimates are informed by state-level data from nationwide weekly surveys (indicated by the black lines and grey boxes). **D and H**: Estimated trend in distributions of time from symptom onset to notification for locally acqu1i0red cases (black line = median; yellow ribbons = 90% distribution quantiles; black dots = time-to-notification of each case). Faded regions indicate where a national trend is used due to low case counts.

#### ii) Successful suppression, re-opening of society

By early April 2020, local case incidence had been driven to very low levels in all Australian states and territories. Substantial numbers of infections continued to be detected in quarantined international arrivals. However, no breaches of quarantine of significant consequence were reported until late May in the state of Victoria [19].

Despite physical distancing measures remaining in place through April, levels of macro-distancing and micro-distancing behaviour steadily waned following peak levels of adherence in the first week of April (Figure 3, panels B, C, F and G). This resulted in a steady increase in estimated transmission potential, although it remained below 1 suggesting that the establishment of community transmission was unlikely throughout this period (Figure 2, panels B and F).

From May through to December 2020, the epidemiology of COVID-19 across Australia was characterised by sustained periods of zero case incidence and intermittent, localised outbreaks (with the exception of the state of Victoria, see below). With the gradual easing of restrictions from May, levels of macro- and micro-distancing behaviour continued to decrease. Accordingly, transmission potential steadily increased and by early June it had exceeded 1 in most states and territories (Figure S1), suggesting that conditions were suitable to sustain onward transmission if there were an undetected importation event or a breakdown in infection control for managed active cases/identified importations.

During the period from late June to mid-October 2020, Australia’s most populous state of New South Wales effectively controlled a series of localised outbreaks (the largest of which involved hundreds of cases). This was achieved during a period where society remained relatively open, though some restrictions on population movement and social gatherings were in place. For example, household and public gatherings were limited to 20 people. Throughout this period, as estimated at the time and now in this retrospective analysis, state-level transmission potential hovered just above 1 (Figure 2, panel B), indicating that levels of population mixing were sufficient to allow escalation of epidemic activity in the general population in the absence of active public health measures to control outbreaks.

We estimate that *R*_eff_ oscillated around 1 throughout this period (Figure 2, panel C). It increased to above 1 at the onset of each incursion and subsequently dropped below 1 as each cluster was contained, with no discernible change in state-level transmission potential (model Component 1) in response to each cluster. These oscillations — strong positive and then negative deviations from the transmission potential — are captured by model Component 2 and are clearly evident in the time-series (Figure 2, panel D). Each of the positive deviations from the transmission potential are consistent with heightened transmission among clusters of cases. Each of the subsequent negative deviations from the transmission potential indicate that the number of offspring from each case of the cluster was fewer than expected given the transmission potential and estimated levels of population mixing. We interpret (and interpreted at the time) this as likely reflecting a strong public health response (*i*.*e*., early detection and isolation of cases associated with the cluster as a result of contact tracing and quarantine). This was consistent with weekly reporting on the performance of contact tracing systems in New South Wales, with 100% of cases interviewed within 24 hours of notification and 100% of close contacts, identified by the case, contacted by public health officials within 48 hours of case notification, from early July through to late October [14].

In mid-November 2020, a sustained period of very low case incidence (*i*.*e*., zero local cases on all but 10 days in the previous 6 months) in the state of South Australia was disrupted by a breach of mandatory quarantine which led to a cluster of more than 20 cases. At the time, society was largely open with only minimal social restrictions in place. We estimate transmission potential to have been 1.71 [95% CrI: 1.47–2.01] as of 14 November in the retrospective analysis (cf. 1.27 [95% CrI: 1.14–1.41] at the time) (Figure S1), suggesting that the risk of establishing an epidemic was reasonably high (relative to the chance of stochastic extinction), and that once established, transmission would be rapid. Supported by our real-time analysis, authorities imposed a strict three-day lockdown across the entire state to enable contact tracers to comprehensively identify and quarantine primary and secondary contacts of cases. Estimated transmission potential declined dramatically around the time of activation of restrictions, and quickly rebounded when restrictions were eased three days later (Figure S1). The incursion was rapidly contained — as result of changes to transmission potential (driven by social restrictions), an effective public health response (*i*.*e*., active case finding and management) and plausibly some favourable stochastic fluctuations — with South Australia returning to zero local case incidence from mid-December 2020.

#### iii) Resurgence of epidemic activity in one large state

In late May 2020, a breach of mandatory quarantine seeded a second epidemic wave in Australia’s second most populous state of Victoria (approximately 6.7 million people). At the time that the epidemic was seeded, many first wave restrictions were still in place. For example, gatherings within households, outdoor spaces, and dining venues were capped at 20 people, and working from home was strongly advised. Transmission potential is estimated to have been 1.07 [95% CrI: 0.88–1.22] at 25 May 2020, suggesting that levels of physical distancing may have been insufficient to prevent escalation of epidemic activity in the general population (Figure 2, panel F).

Furthermore, from the earliest stages of the epidemic, our model estimated a strong positive deviation from the transmission potential (Component 2 positively biased, Figure 2, panel H), corresponding to an estimate for the *R*_eff_ *>* 1 (95% chance of *R*_eff_ exceeding 1 by 1 June 2020 in the retrospective analysis) reflecting heightened transmission. Demographic and socio-economic assessments of the outbreak [20, 21, 22] showed that early affected areas had higher than average household sizes and a large proportion of essential and casualised workers who were unable to work from home. Thus our model findings concurred with the observed epidemiological characteristics — that the virus was predominantly spreading in subsections of the population with higher-than-average rates of social contact — and supported public health decision making at the time.

By 1 July 2020, there were more than 600 active cases and 129 newly reported cases with an estimated *R*_eff_ of 1.33 [95% CrI: 1.25–1.41] (Figure 2, panel G). From 9 July 2020, stay-at-home policies (denoted Stage 3 restrictions) were reinstated across metropolitan Melbourne. Despite these policies, the epidemic continued to grow through July, reaching a peak of 446 daily cases by date of symptom onset on 24 July 2020. More severe stay-at-home restrictions (denoted Stage 4) were enacted in metropolitan Melbourne on 2 August, including a night-time curfew, restrictions on movement more than 5km from a person’s residence, and stricter definitions of essential workers and businesses including invigilation of a work permit requirement.

During the periods of Stage 3 and 4 restrictions, we observed strong increases in macro- and micro-distancing behaviour, which was reflected by a decrease in state-level transmission potential from around 1 in early June to a minimum of 0.72 [95% CrI: 0.62–0.86] on 23 August 2020 (Figure 2, panel F), two weeks after the implementation of Stage 4 restrictions.

Following an initial sharp rise in the *R*_eff_ from well below 1 in mid-May to a peak of 1.61 [95% CrI: 1.46–1.79] at 14 June 2020, the *R*_eff_ steadily decreased over the next eight weeks (Figure 2, panel G). We estimate that *R*_eff_ fell below the critical threshold of 1 on 25 July, approximately one week prior to the implementation of Stage 4 restrictions. With Stage 4 restrictions in place, *R*_eff_ settled between 0.8 and 1 for another eight weeks.

While both transmission potential and *R*_eff_ declined over this period, we estimated *R*_eff_ to be consistently higher than transmission potential (*i*.*e*., there was a strong positive deviation in Component 2) reflecting persistent transmission in subsections of the population with higher-than-average rates of social contact. This was consistent with other epidemiological assessments of the outbreak which suggested that transmission was concentrated in populations that were less able to physically distance (*e*.*g*., healthcare workers, residents of aged care facilities, meat workers public housing residents) [20, 21, 22]. A substantial proportion of cases were in health-care workers and aged care facilities, particularly during the tail of the epidemic. Each of these settings required specifically targeted interventions to bring transmission under control, which were distinct from the impacts of population level measures. This may partly explain why transmission persisted for many weeks when severe stay-at-home restrictions were active, since these measures primarily target transmission in the broader community and are logically less effective at controlling transmission in essential workplaces and institutional settings.

Definitive control of the epidemic was achieved by early November 2020, when zero local case incidence reported in Victoria for the first time since April 2020.

The pattern in Component 2 for Victoria, where it deviated strongly above zero in the earliest stages of the epidemic, persisted above zero for many months, and returned to around zero once the epidemic was definitely contained, is in contrast to the oscillations seen in New South Wales from June to October.

## Discussion

We have presented a novel semi-mechanistic modelling framework for assessing transmissibility of SARS-CoV-2 from periods of high to low — or zero — case incidence, with a seamless and coherent transition in interpretation across the changing epidemiological situations. Using time-series data on cases and population behaviours, our model computes three metrics within a single framework: the effective reproduction number for active cases (*R*_eff_), the population-wide transmission potential (TP), and the deviation between *R*_eff_ and TP (C2). Our model has been applied (in real-time) to Australian data throughout the pandemic and continues to support the public health response. Here, our analysis of the first 12 months of the pandemic has demonstrated how these quantities enable the tracking and planning of progress towards the control of large outbreaks (as seen in Victoria), maintenance of virus suppression (as seen in New South Wales), and monitoring the risk posed by re-introduction of the virus (as seen in South Australia).

Our approach addresses a major challenge in epidemic situational awareness by enabling assessment of epidemic risk — via the TP — when cases are driven to low levels or (temporary) elimination is achieved. During periods of viral transmission, the model also provides new insight into epidemic dynamics via the deviation between *R*_eff_ and TP (C2). Further, the TP provides near-real-time assessment of trends in population macro- and micro-distancing behaviours that fluctuate in response to changing social restrictions, risk perception, and other factors such as school holidays. In combination, knowledge gained from *R*_eff_, TP and C2 enables policymakers to monitor the relative impacts of community-wide social restrictions and consider the need for more targeted response measures [13].

Social and behavioural data have been used extensively in other countries to support COVID-19 situational assessment [1, 23, 24, 25, 26, 27]. In the UK, the CoMix study [23] has been collecting contact data on a fortnightly basis since March 2020 and reporting “*R*_*c*_” (the basic reproduction number under control measures), to the UK government’s Scientific Pandemic Influenza Group on Modelling, Operational sub-group (SPI-M-O). Conceptually, CoMix’s *R*_*c*_ is akin to our TP. However, by synthesising behavioural data from multiple sources, accounting for both micro- and macro-distancing behaviours (thus estimating ‘effective’ contacts), and incorporating the effect of case surveillance, our approach is likely to capture a more complete picture of the population-wide potential for virus transmission. Further, by estimating TP and *R*_eff_ within the same modelling framework (and thus computing C2), our analysis provides a richer and more coherent epidemiological interpretation than that offered through independent measurement and reporting of each metric. Our case studies demonstrate how this richness has supported (and continues to support) the Australian COVID-19 response.

Despite its demonstrated impact, there are limitations to our approach. Firstly, it relies on data from frequent, population-wide surveys. In Australia, these data are collected for government and made available to our analysis team by a market research company which has access to an established “panel” of individuals who have agreed to take part in surveys of public opinion. Researchers and governments in many other countries have used such companies for rapid data collection to support pandemic response [23, 25]. However, these survey platforms are not readily available in all settings. Further, the sampling strategy did not allow for surveying individuals without internet access, low literacy or limited English language skills, or communication or cognitive difficulties. Further, individuals under 18 years of age were not represented in our surveys. Nor were these survey results available for the pre-pandemic period, limiting our ability to estimate what a true behavioural baseline would be for the Australian population.

While the patterns of TP, *R*_eff_ and C2 observed over time in Australia are consistent with “in field” epidemiological assessments, and while the methods have demonstrated impact in supporting decision making, a direct quantification of the validity of the TP is not straightforward. For example, whether self-reported adherence to the 1.5 m rule is a reliable covariate for change in the per contact probability of transmission over time is difficult to assess. If transmission were to become widespread in Australia; and therefore cases become more representative of the general population rather than specific subsets, *R*_eff_ and TP estimates would be expected to converge. However in the absence of such a natural experiment, no ground truth for this unobserved parameter exists with which to quantitatively validate the model calibration.

In Australia, our methods are not only embedded in state and national situational assessment [14] but also national response planning. Since the model incorporates a mechanistic understanding of the impacts of physical distancing behaviour on both household and non-household transmission, it can therefore be used to predict the impact of interventions on actual and potential transmission [16].

Unlike other approaches that make assumptions about impacts of different interventions on behaviour, we directly measure and account for behavioural responses, providing a much more proximal way of assessing the effects of interventions [28]. Further, while detailed data on the demographics and transmission settings for cases in Australia is unavailable, our method considers deviation (the C2) from the regional average (the TP). It is therefore less susceptible to conflation between an epidemic stochastically moving between settings of different transmissibility, and changes in population-wide transmission potential.

While not addressed in this article, our semi-mechanistic model structure enables us to perform independent estimates of the relative transmissibility of variants compared to ancestral strains. In doing so, we account for variability in the types of contacts made when low restrictions are applied [3]. We are able to estimate differences between variants in the probability of transmission per unit of contact-time, for example from detailed attack rate data from overseas. These probabilities can then be combined with our estimates from Australian case data to adjust our estimates of TP under different levels of restrictions for current and emerging variants. We have also recently updated our modelling framework to account for the effects of vaccination on the TP (reported in the Australian Government’s Common Operating Picture from 27 August 2021 [14]). This enables us to consider the effect of varying levels of population vaccination coverage, age-based vaccination prioritisation strategies, and levels of restrictions on the ability of the Delta variant (and future possible variants) to spread in the population. These analyses underpin the recent Australian national COVID-19 re-opening plan [16] and will be reported elsewhere.

Our novel methods provide new insight into epidemic dynamics in both low and high incidence settings. The analyses have become an indispensable tool supporting the Australian COVID-19 response, through both situational assessment and strategic planning processes.

## Materials and Methods

### Model overview

We estimate the time-varying ability of SARS-CoV-2 to spread in a population using a novel semi-mechanistic model informed by data on cases, population behaviours and health system effectiveness. We separately model transmission from locally acquired cases (local-to-local transmission) and from overseas acquired cases (import-to-local transmission). We model local-to-local transmission (*R*_eff_) using two components:

1. the average population-level trend in transmissibility driven by interventions that primarily target transmission from local cases, specifically changes in physical distancing behaviour and case targeted measures (Component 1); and
2. short-term fluctuations in *R*_eff_ to capture stochastic dynamics of transmission, such as clusters of cases and short periods of lower-than-expected transmission, and other factors factors influencing *R*_eff_ that are otherwise unaccounted for by the model (Component 2).

During times of disease activity, Components 1 and 2 are combined to provide an estimate of the local *R*_eff_ as traditionally measured. In the absence of disease activity, Component 1 is interpreted as the potential for the virus, if it were present, to establish and maintain community transmission (*>* 1) or otherwise (*<* 1).

### Case data

We used line-lists of reported cases for each Australian state and territory extracted from the Australian National Notifiable Disease Surveillance System (NNDSS). The line-lists contain the date when the individual first exhibited symptoms, date when the case notification was received by the jurisdictional health department and where the infection was acquired (*i*.*e*., overseas or locally).

### Modelling the impact of physical distancing

#### Overview

To investigate the impact of distancing measures on SARS-CoV-2 transmission, we distinguish between two types of distancing behaviour: 1) macro-distancing *i*.*e*., reduction in the rate of non-household contacts; and 2) micro-distancing *i*.*e*., reduction in transmission probability per non-household contact.

We used data from nationwide surveys to estimate trends in specific macro-distancing (average daily number of non-household contacts) and micro-distancing (proportion of the population always keeping 1.5 metre physical distance from non-household contacts) behaviours over time. We used these survey data to infer state-level trends in macro- and micro-distancing behaviour over time, with additional information drawn from trends in mobility data.

### Estimating changes in macro-distancing behaviour

To estimate trends in macro-distancing behaviour, we used data from: two waves of a national survey conducted in early April and early May 2020 by the University of Melbourne; and weekly waves of a national survey conducted by the Australian government from late May 2020. Respondents were asked to report the number of individuals that they had contact with outside of their household in the previous 24 hours. Note that the first wave of the University of Melbourne survey was fielded four days after Australia’s most intensive physical distancing measures were recommended nationally on 29 March 2020.

Given these data, we used a statistical model to infer a continuous trend in macro-distancing behaviour over time. This model assumed that the daily number of non-household contacts is proportional to a weighted average of time spent at different types of location, as measured by Google mobility data. The five types of places are: parks and public spaces; residential properties; retail and recreation; public transport stations; and workplaces. We fit a statistical model that infers the proportion of non-household contacts occurring in each of these types of places from:

- a survey of location-specific contact rates pre-COVID-19 [29]; and
- a separate statistical model fit to the national average numbers of non-household contacts from a pre-COVID-19 contact survey and contact surveys fielded post-implementation of COVID-19 restrictions.

Waning in macro-distancing behaviour is therefore driven by Google mobility data on increasing time spent in each of the different types of locations since the peak of macro-distancing behaviour.

### Estimating changes in micro-distancing behaviour

To estimate trends in micro-distancing behaviour, we used data from weekly national surveys (first wave from 27–30 March 2020) to assess changes in behaviour in response to COVID-19 public health measures. Respondents were asked to respond to the question: ‘Are you staying 1.5m away from people who are not members of your household’ on a five point scale with response options “No”, “Rarely”, “Sometimes”, “Often” and “Always”.

These behavioural survey data were used in a statistical model to infer the trend in micro-distancing behaviour over time. Micro-distancing behaviour was assumed to be non-existent prior to the first epidemic wave of COVID-19, and the increase in micro-distancing behaviour to its peak was assumed to follow the same trend as macro-distancing behaviour — implying that the population simultaneously adopted both macro- and micro-distancing behaviours around the times that restrictions were implemented. The behavioural survey data was then used to infer the date of peak micro-distancing behaviour (assumed to be the same in all states), the proportion of the population adopting micro-distancing behaviour, and the rate at which micro-distancing behaviour is waning from that peak in each state.

### Incorporating estimated changes in distancing behaviour in the model of transmission potential

These state-level macro-distancing and micro-distancing trends were then used in the model of transmission potential to inform the reduction in non-household transmission rates. Since the macro-distancing trend is calibrated against the number of non-household contacts, the rate of non-household transmission scales directly with this inferred trend. The probability of transmission per non-household contact is assumed to be proportional to the fraction of survey participants who report that they always maintain 1.5m physical distance from non-household contacts. The constant of proportionality is estimated in the model of transmission potential.

The estimated rate of waning of micro-distancing is sensitive to the metric used. If a different metric of micro-distancing (*e*.*g*., the fraction of respondents practicing good hand hygiene) were used, this might affect the inferred rate of waning of micro-distancing behaviour, and therefore increasing the transmission potential.

### Modelling the impact of quarantine of overseas arrivals

We model the impact of quarantine of overseas arrivals via a ‘step function’ reflecting three different quarantine policies: self-quarantine of overseas arrivals from specific countries prior to March 15; self-quarantine of all overseas arrivals from March 15 up to March 27; and mandatory quarantine of all overseas arrivals after March 27 (Figure S8). We make no prior assumptions about the effectiveness of quarantine at reducing *R*_eff_ import, except that each successive change in policy increased that effectiveness.

### Model limitations

While we had access to data on whether cases are locally acquired or overseas acquired, no data were available on whether each of the locally acquired cases were infected by an imported case or by another locally acquired case. This data would allow us to disentangle the two transmission rates. Without this data, we can separate the denominators (number of infectious cases), but not the numerators (number of newly infected cases) in each group at each point in time. With access to such data, our method could provide more precise estimates of *R*_eff_.

### Model description

We developed a semi-mechanistic Bayesian statistical model to estimate *R*_eff_, or *R*(*t*) hereafter, the effective rate of transmission of of SARS-CoV-2 over time, whilst simultaneously quantifying the impacts on *R*(*t*) of a range of policy measures introduced at national and regional levels in Australia.

#### Observation model

A straightforward observation model to relate case counts to the rate of transmission is to assume that the number of new locally-acquired cases 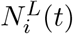 at time *t* in region *i* is (conditional on its expectation) Poisson-distributed with mean *λ*_*i*_(*t*) given by the product of the total infectiousness of infected individuals *I*_*i*_(*t*) and the time-varying reproduction rate *R*_*i*_(*t*):

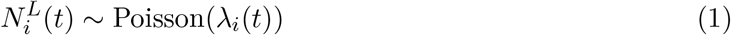

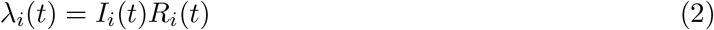

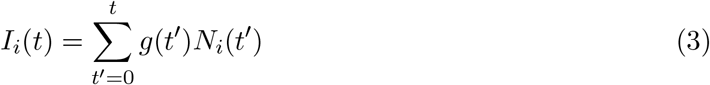

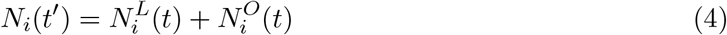

where the total infectiousness, *I*_*i*_(*t*), is the sum of all active infections *N*_*i*_(*t′*) — both locally-acquired 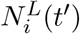 and overseas-acquired 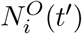 — initiated at times *t′* prior to *t*, each weighted by an infectivity function *g*(*t′*) giving the proportion of new infections that occur *t′* days post-infection. The function *g*(*t′*) is the probability of an infector-infectee pair occurring *t′* days after the infector’s exposure, *i*.*e*., a discretisation of the probability distribution function corresponding to the generation interval.

This observation model forms the basis of the maximum-likelihood method proposed by White and Pagano (2007) [12] and the variations of that method by Cori et al. (2013) [9], Thompson et al. (2019) [10] and Abbott et al. (2020) [30] that have previously been used to estimate time-varying SARS-CoV-2 reproduction numbers in Australia [18].

We extend this model to consider separate reproduction rates for two groups of infectious cases, in order to model the effects of different interventions targeted at each group: those with locally-acquired cases 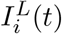, and those with overseas acquired cases 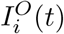, with corresponding reproduction rates 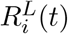 and 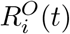. These respectively are the rates of transmission from imported cases to locals, and from locally-acquired cases to locals. We also model daily case counts as arising from a Negative Binomial distribution rather than a Poisson distribution to account for potential clustering of new infections on the same day, and use a state- and time-varying generation interval distribution *g*_*i*_(*t′, t′*) (detailed in *Surveillance effect model*):

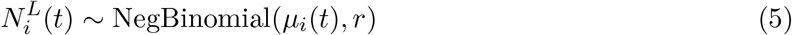

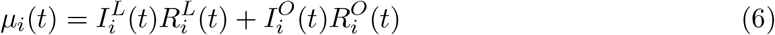

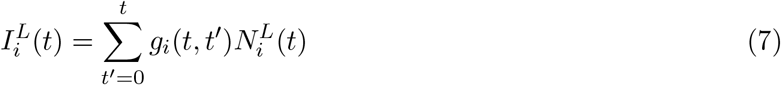

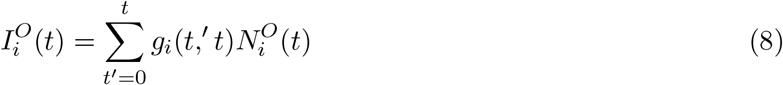

where the negative binomial distribution is parameterised in terms of its mean *μ*_*i*_(*t*) and dispersion parameter *r*. In the commonly used probability and dispersion parameterisation with probability *ψ* the mean is given by *μ* = *ψr/*(1 − *ψ*).

Note that if data were available on the whether the source of infection for each locally-acquired case was another locally-acquired case or an overseas-acquired cases, we could split this into two separate analyses using the observation model above; one for each transmission source. In the absence of such data, the fractions of all transmission attributed to sources of each type is implicitly inferred by the model, with an associated increase in parameter uncertainty.

We provide the model with additional information on the rate of import-to-local transmission by adding a further likelihood term to the model for known events of import-to-local transmission since the implementation of mandatory hotel quarantine:

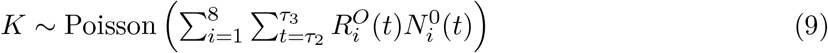

where *K* is the total number of known events of transmission *from* overseas-acquired cases occurring within Australia from *τ*_2_ = 2020-03-28 to *τ*_3_ = 2020-12-31. These events are largely transmission events within hotel quarantine facilities, some of which led to outbreaks of local-to-local transmission. Prior to this period, import-to-local transmission events cannot be reliably distinguished from local-to-local transmission events.

When estimating *R*_eff_ from recent case count data, care must be taken to account for under-reporting of recent cases (those which have yet to be detected), because failing to account for this under-reporting can lead to estimates of *R*_eff_ that are biased downwards. We correct for this right-truncation effect by first estimating the fraction of locally-acquired cases on each date that we would expect to have detected by the time the model is run (detection probability), and correcting both the infectiousness terms 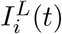, and the observed number of new cases 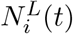. We calculate the detection probability for each day in the past from the empirical cumulative distribution function of delays from assumed date of infection to date of detection over a recent period (see *Surveillance effect model*). We correct the infectiousness estimates 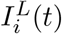 by dividing the number of newly infected cases on each day 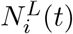 by this detection probability — to obtain the expected number of new infections per day — before summing across infectiousness. We correct the observed number of new infections by a modification to the negative binomial likelihood; multiplying the expected number of cases by the detection probability to obtain the expected number of cases observed in the (uncorrected) time series of locally-acquired cases.

#### Reproduction rate models

We model the onward reproduction rates for overseas-acquired and locally-acquired cases in a semi-mechanistic way. Reproduction rates for local-to-local transmission are modelled as a combination of a deterministic model of the population-wide transmission potential for that type of case, and a correlated time series of random effects to represent stochastic fluctuations in the reporting rate in each state over time. Import-to-local transmission is modelled in a mechanistic way:

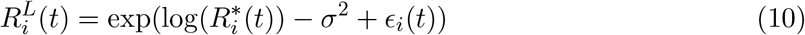

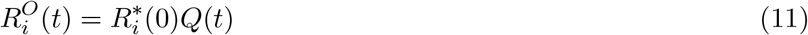

For locally-acquired cases, the state-wide average transmission rate at time 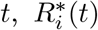, is given by a deterministic epidemiological model of population-wide transmission potential that considers the effects of distancing behaviours. The correlated time series of random effects *ϵ*_*i*_(*t*) represents stochastic fluctuations in these local-local reproduction rates in each state over time — for example due to clusters of transmission in sub-populations with higher or lower reproduction rates than the general population. We consider that the transmission potential 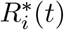 is the average of individual reproduction rates over the entire state population, whereas the effective reproduction number 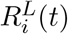 is the average of individual reproduction rates among a (non-random) sample of individuals – those that make up the active cases at that point in time. We therefore expect that the long-term average of 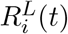 will equate to 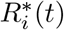. The relationship between these two is therefore defined such that the hierarchical distribution over 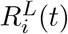 is marginally (with respect to time) a log-normal distribution with mean 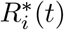. The parameter *σ*^2^ is the marginal variance of the *ϵ*_*i*_, as defined in the kernel function of the Gaussian process.

For overseas-acquired cases the population-wide transmission rate at time 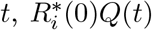, is the baseline rate of transmission (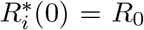 ; local-to-local transmission potential in the absence of distancing behaviour or other mitigation) multiplied by a quarantine effect model, *Q*(*t*), that encodes the efficacy of the three different overseas quarantine policies implemented in Australia (described below).

We model 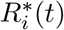, the population-wide rate of local-to-local transmission at time *t*, as the sum of two components: the rate of transmission to members of the same household, and to members of other households. Each of these components is computed as the product of the number of contacts, and the probability of transmission per contact. The transmission probability is in turn modelled as a binomial process considering the duration of contact with each person and the probability of transmission per unit time of contact. This mechanistic consideration of the contact process enables us to separately quantify how macro- and micro-distancing behaviours impact on transmission, and to make use of various ancillary measures of both forms of distancing:

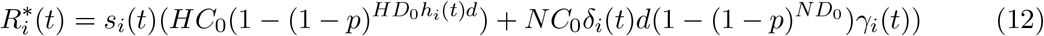

where: *s*(*t*) is the effect of surveillance on transmission, due to the detection and isolation of cases (detailed below); *HC*_0_ and *NC*_0_ are the baseline (*i*.*e*., before adoption of distancing behaviours) daily rates of contact with, respectively, people who are, and are not, members of the same household; *HD*_0_ and *ND*_0_ are the baseline average total daily duration of contacts with household and non-household members (measured in hours); *d* is the average duration of infectiousness in days; *p* is the probability of transmitting the disease per hour of contact, and; *h*_*i*_(*t*), *δ*_*i*_(*t*), *γ*_*i*_(*t*) are time-varying indices of change relative to baseline of the duration of household contacts, the number of non-household contacts, and the transmission probability per non-household contact, respectively (modifying both the duration and transmission probability per unit time for non-household contacts).

The first component in Equation (12) is the rate of household transmission, and the second is the rate of non-household transmission. Note that the duration of infectiousness *d* is considered differently in each of these components. For household members, the daily number of household contacts is typically close to the total number of household members, hence the expected number of household transmissions saturates at the household size; so the number of days of infectiousness contributes to the probability of transmission to each of those household members. This is unlikely to be the case for non-household members, where each day’s non-household contacts may overlap, but are unlikely to be from a small finite pool. This assumption would be unnecessary if contact data were collected on a similar timescale to the duration of infectiousness, though issues with participant recall in contact surveys mean that such data are unavailable.

The parameters *HC*_0_, *HD*_0_, and *ND*_0_ are all estimated from a contact survey conducted in Melbourne in 2015 [29]. *NC*_0_ is computed from an estimate of the total number of contacts per day for adults from [31], minus the estimated rate of household contacts. Whilst [29] also provides an estimate of the rate of non-household contacts, the method of data collection (a combination of ‘individual’ and ‘group’ contacts) makes it less comparable with contemporary survey data than the estimate of [31].

The expected duration of infectiousness *d* is computed as the mean of the non-time-varying discrete generation interval distribution:

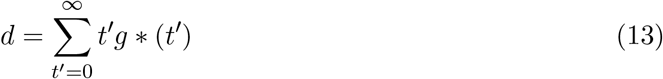

and change in the duration of household contacts over time *h*_*i*_(*t*) is assumed to be equivalent to change in time spent in residential locations in region *i*, as estimated by the mobility model for the data stream *Google: time at residential*. In other words, the total duration of time in contact with household members is assumed to be directly proportional to the amount of time spent at home. Unlike the effect on non-household transmission, an increase in macro-distancing is expected to slightly increase household transmission due to this increased contact duration.

The time-varying parameters *δ*_*i*_(*t*) and *γ*_*i*_(*t*) respectively represent macro- and micro-distancing; behavioural changes that reduce mixing with non-household members, and the probability of transmission for each of non-household member contact. We model each of these components, informed by population mobility estimates from the mobility model and calibrated against data from nationwide surveys of contact behaviour.

#### Surveillance effect model

Disease surveillance — both screening of people with COVID-like symptoms and performing contact tracing — can improve COVID-19 control by placing cases in isolation so that they are less likely to transmit the pathogen to other people. Improvements in disease surveillance can therefore lead to a reduction in transmission potential by isolating cases more quickly, and reducing the time they are infectious but not isolated. Such an improvement changes two quantities: the population average transmission potential *R*^*^(*t*) is reduced by a factor *s*_*i*_(*t*); and the generation interval distribution *g*(*t, t′*) is shortened, as any transmission events are more likely to occur prior to isolation.

We model both of these functions using a region- and time-varying estimate of the discrete probability distribution over times from infection to detection *f*_*i*_(*t, t′*):

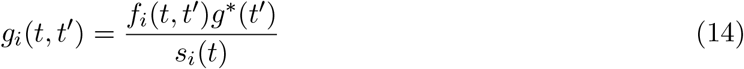

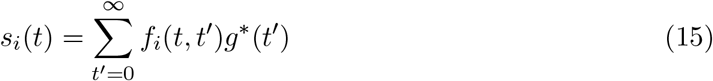

where *g*^*^(*t′*) is the baseline generation interval distribution, representing times to infection in the absence of detection and isolation of cases, *s*_*i*_(*t*) is a normalising factor — and also the effect of surveillance on transmission — and *f*_*i*_(*t, t′*) is a region- and time-varying probability density over periods from infection to isolation *t′*. In states/territories and at times when cases are rapidly found and placed in isolation, the distribution encoded by *f*_*i*_(*t, t′*) has most of its mass on small delays, average generation intervals are shortened, and the surveillance effect *s*_*i*_(*t*) tends toward 0 (a reduction in transmission). At times when cases are not found and isolated until after most of their infectious period has passed, *f*_*i*_(*t, t′*) has most of its mass on large delays, generation intervals are longer on average, and *s*_*i*_(*t*) tends toward 1 (no effect of reduced transmission).

We model the region- and time-varying distributions *f*_*i*_(*t, t′*) empirically via a time-series of empirical distribution functions computed from all observed infection-to-isolation periods observed within an adaptive moving window around each time *t*. Since dates of infection and isolation are not routinely recorded in the dataset analysed, we use 5 days prior to the date of symptom onset to be the assumed date of infection, and the date of case notification to be the assumed date of isolation. This will overestimate the time to isolation and therefore underestimate the effect of surveillance when a significant proportion of cases are placed into isolation prior to testing positive — *e*.*g*., during the tail of an outbreak being successfully controlled by contact tracing.

For a given date and state/territory, the empirical distribution of delays from symptom onset to notification is computed from cases with symptom onset falling within a time window around that date, with the window selected to be the smallest that will yield at least 500 observations; but constrained to between one and eight weeks.

Where a state/territory does not have sufficient cases to reliably estimate this distribution in an eight week period, a national estimate is used instead. Specifically, if fewer than 100 cases, the national estimate is used, if more than 500 the state estimate is used, and if between 100 and 500 the distribution is a weighted average of state and national estimates.

The national estimate is obtained via the same method but with no upper limit on the window size and excluding data from Victoria since 14 June, since the situation during the Victorian outbreak after this time is not likely to be representative of surveillance in states with few cases.

#### Macro-distancing model

The population-wide average daily number of non-household contacts at a given time can be directly estimated using a contact survey. We therefore used data from a series of contact surveys commencing immediately after the introduction of distancing restrictions to estimate *δ*_*i*_(*t*) independently of case data. To infer a continuous trend of *δ*_*i*_(*t*), we model the numbers of non-household contacts at a given time as a function of mobility metrics considered in the mobility model. We model the log of the average number of contacts on each day as a linear model of the log of the ratio on baseline of five Google metrics of time spent at different types of location: residential, transit stations, parks, workplaces, and retail and recreation:

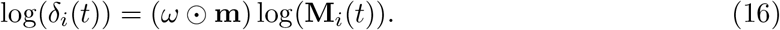

where *ω* is the the vector of 5 coefficients, **m** is an vector of length 5 containing of ones, except for the element corresponding to time at residential locations, which has value 1, and ⊙ indicates the elementwise product. This constrains the direction of the effect of increasing time spent at each of these locations to be positive (more contacts), except for time at residential, which we constrain to be negative. The intercept of the linear model (average daily contacts at baseline) is given an prior formed from the daily number of non-household contacts in a pre-COVID-19 contact survey [29]. Since our aim is to capture general trends in mobility rather than daily effects, we model the weekly average of the daily number of contacts, by using smoothed estimates of the Google mobility metrics.

Whilst we aim to model weekly rather than daily variation in contact rates, when fitting the model to survey data we account for variation among responses by day of the week by modelling the fraction of the weekly number of contacts falling on each day of the week (the length-seven vector in each state and time **D**_*i*_(*t*)) and using this to adjust the expected number of contacts for each respondent based on the day of the week they completed the survey. To account for how the weekly distribution of contacts has changed over time as a function of mixing restrictions (*e*.*g*., a lower proportion of contacts on weekdays during periods when stay-at-home orders were in place) we model the weekly distribution of contacts itself as a function of deviation in the weekly average of the daily number of contacts, with length-seven vector parameters *α* and *θ*. We use the softmax (normalised exponential) function to transform this distribution to sum to one, then multiply the resulting proportion by 7 to reweight the weekly average daily contact rate to the relevant day of the week.

Combining the baseline average daily contact rate *NC*_0_, mobility-driven modelled change in contact rates over time *δ*_*i*_(*t*), and time-varying day of the week effects **D**_*i*_(*t*) we obtain an expected number of daily contacts for each survey response *NC*_*k*_:

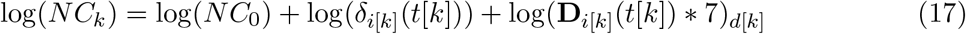

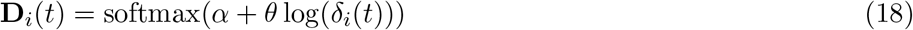

where *i*[*k*], *t*[*k*], and *d*[*k*] respectively indicate the state, time, and day of the week on which respondent *k* filled in the survey.

We model the number of contacts from each survey respondent as a draw from an interval-censored discrete lognormal distribution. This choice of distribution enables us to account for the *ad-hoc* rounding of reported numbers of contacts (responses larger than 10 tend to be ‘heaped’ on multiples of 10 and 100), whilst also accounting for heavy upper tail in numbers of reported contacts. The support of this distribution is the integers from 0 to 10 inclusive, and the intervals 11-20, 21-50, and 50-999. Reported daily contact rates ≥1000 are excluded as these are considered implausible for our definition of a contact. The probability mass function of this distribution is the integral across these ranges of a lognormal distribution with parameters *μ*_*k*_ and *τ*, parameterised such that the mean of the distribution is *NC*_*k*_:

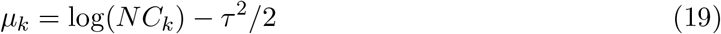

#### Micro-distancing model

Unlike with macro-distancing behaviour and contact rates, there is no simple mathematical framework linking change in micro-distancing behaviours to changes in non-household transmission probabilities. We must therefore estimate the effect of micro-distancing behaviour on transmission via case data. We implicitly assume that any reduction in local-to-local transmission potential that is not explained by changes to the numbers of non-household contacts, the duration of household contacts, or improved disease surveillance is explained by the effect of micro-distancing on non-household transmission probabilities.

Whilst it is not necessary to use ancillary data to estimate the effect that micro-distancing has at its peak, we use behavioural survey data to estimate the temporal trend in micro-distancing behaviour, in order to estimate to what extent adoption of that behaviour has waned and how that has affected transmission potential.

We therefore model *γ*_*t*_ (a time-varying index of change relative to baseline of transmission probability per non-household contact, see Equation (12)), as a function of the proportion of the population adhering to micro-distancing behaviours. We consider adherence to the ‘1.5m rule’ as indicative of this broader suite of behaviours due to the availability of data on this behaviour in a series of weekly behavioural surveys beginning prior to the last distancing restriction being implemented [32]. We consider the number 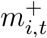 of respondents in region *i* on survey wave commencing at time *t* replying that they ‘always’ keep 1.5m distance from non-household members, as a binomial sample with sample size *m*_*i,t*_. We use a generalised additive model to estimate *c*_*i*_(*t*), the proportion of the population in region *i* responding that they always comply as a the intervention stage, smoothed over time. Intervention stages are defined as periods of a continuous state of stay-at-home order, and this state thus switches each time a stay-at-home order is started, ended, or significantly changed. This state switching allows the model to react to sudden changes in compliance behaviour when orders are made or rescinded. We assume that the temporal pattern in the initial rate of adoption of the behaviour is the same as for macro-distancing behaviours — the adoption curve estimated from the mobility model. In other words, we assume that all macro- and micro-distancing behaviours were adopted simultaneously around the time the first population-wide restrictions were put in place in March and April 2020. However we do not assume that these behaviours peaked at the same time or subsequently followed the same temporal trend. The model for the proportion complying with this behaviour is therefore:

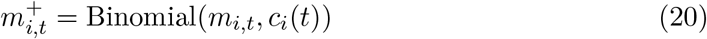

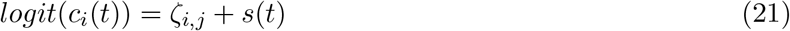

where *ζ*_*i,j*_ is intervention state *j* in region *i*, and *s* is a smoothing function over time *t*.

Given *c*_*i*_(*t*), we model *γ*_*i*_(*t*) as a function of the degree of micro-distancing relative to the peak:

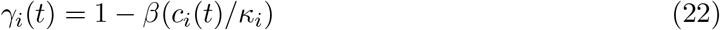

where *κ*_*i*_ is the peak of compliance, or maximum of *c*_*i*_(*t*), and *β* is inferred from case data in the main *R*_eff_ model.

#### Overseas quarantine model

We model the effect of overseas quarantine *Q*(*t*) via a monotone decreasing step function with values constrained to the unit interval, and with steps at the known dates *τ*_1_ and *τ*_2_ of changes in quarantine policy:

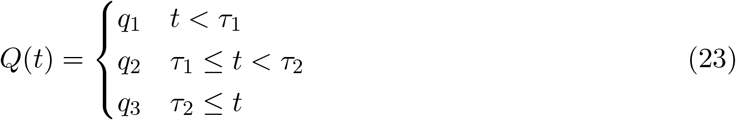

where *q*_1_ *> q*_2_ *> q*_3_ and all parameters are constrained to the unit interval.

#### Error models

The correlated time-series of deviance between transmission potential the effective reproduction rate for local-to-local transmission in each region *ϵ*_*i*_(*t*) is modelled as a zero-mean Gaussian process (GP) with covariance structure reflecting temporal correlation in errors within each region, but independent between regions. We use a Matern 5/2 covariance function *k*, enabling a mixture of relatively smooth trends and local ‘roughness’ to represent the sudden rapid growth of cases that can occur with a high-transmission cluster. Kernel parameters *σ* and *l* are the same across regions:

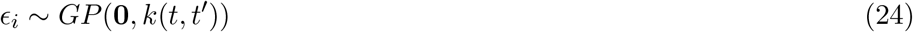

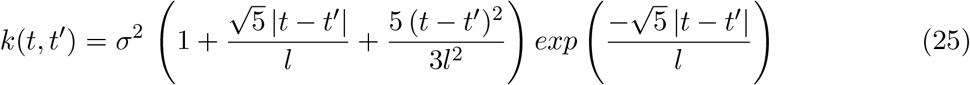

#### Components of local transmission potential

We model the rate of transmission from locally acquired cases as a combination of the time-varying mechanistic model of transmission rates 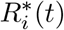, and a temporally-correlated error term 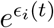. This structure enables inference of mechanistically interpretable parameters whilst also ensuring that statistical properties of the observed data are represented by the model. Moreover, these two parts of the model can also be interpreted in epidemiological terms as two different components of transmission rates:

1. **Component 1 (TP)** – transmission rates averaged over the whole state population, representing how macro- and micro-distancing, and other factors affect the potential for widespread community transmission 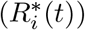, and
2. **Component 2 (C2)** – the degree to which the transmission rates of the population of current active cases deviates from the average statewide transmission rate 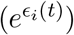.

Component 2 reflects the fact that the population of current active cases in each state at a given time will not be representative of the the state-wide population, and may be either higher (*e*.*g*., when cases arise from a cluster in a high-transmission environment) or lower (*e*.*g*., when clusters are brought under control and cases placed in isolation).

Component 1 (TP) can therefore be interpreted as the expected rate of transmission if cases were widespread (population-representative) in the community. The product of Components 1 and 2 (*R*_eff_) can be interpreted as the rate of transmission in the sub-population making up active cases at a given time.

Where a state has active cases in one or more clusters, the combination of these components gives the apparent rate of transmission in those clusters (*R*_eff_), given by Equation 10. This reflects the interpretation that TP captures the population mean of a distribution over individual-level reproduction numbers, and *R*_eff_ is the mean of a (non-random) sample from that distribution — the population comprising cases at that point in time. While not used in the public health context in Australia, the epidemiological interpretation of the *R*_eff_ when a state has no active cases is the rate of spread expected if an index case were to occur in a random sub-population. Because the amplitude of this error term is learned from the data, this is informative as to the range of plausible rates of spread that might be expected from a case being introduced into a random sub-population. However, the mean of this distribution, TP, may play a similar role and has proven to be a more interpretable quantity for end users of this model.

#### Parameter values and prior distributions

Tables S2 and S4 give the prior distributions of parameters in the semi-mechanistic and time-series (*ϵ*) parts of the model respectively. Table S3 gives fixed parameter values used in the semi-mechanistic part of the model.

The parameters of the generation interval distribution are the posterior mean parameter estimates corresponding to a lognormal distribution over the serial interval estimated by [33]. The shape of the generation interval distribution for SARS-CoV-2 in comparable populations is not well understood, and a number of alternative distributions have been suggested by other analyses. A sensitivity analysis performed by running the model with alternative generation interval distributions (not presented here) showed that parameter estimates were fairly consistent between these scenarios, and the main findings were unaffected. A full, formal analysis of sensitivity to this and other assumptions will be presented in a future publication.

No ancillary data are available to inform *p*, the probability of transmission per hour of contact in the absence of distancing behaviour. However, at *t* = 0, holding *HC*_0_, *NC*_0_ *HD*_0_, and *ND*_0_ constant, there is a deterministic relationship between *p* and 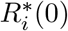 (the basic reproduction rate, which is the same for all states). The parameter *p* is therefore identifiable from transmission rates at the beginning of the first epidemic wave in Australia. We define a prior on *p* that corresponds to a prior over 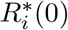 matching the averages of the posterior means and 95% credible intervals for 11 European countries as estimated by [28] in a sensitivity analysis where the mean generation interval was 5 days — similar to the serial interval distribution assumed here. This corresponds to a prior mean of 2.79, and a standard deviation of 1.70 for 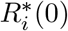. This prior distribution over *p* was determined by a Monte-Carlo moment-matching algorithm, integrating over the prior values for *HC*_0_, *NC*_0_ *HD*_0_, and *ND*_0_.

#### Model fitting

We fitted (separate) models of *c*_*i*_(*t*) and *NC*_0_*δ*_*i*_(*t*) to survey data alone in order to infer trends in those parameters as informed by survey data. These are shown in Figures S5–S6. We used the posterior means of each of these model outputs as inputs into the *R*_eff_ model. The posterior variance of each of these quantities is largely consistent over time and between states, and the absolute effect of each is scaled by other parameters (e.g. *β*), meaning that uncertainty in these quantities is largely not identifiable from uncertainty in other scaling parameters. As a consequence, propagation of uncertainty in these parameters into the *R*_eff_ model (as was performed in a previous iteration of the model) has little impact on estimates of *R*_eff_ and transmission potential, so is avoided for computational brevity.

Inference was performed by Hamiltonian Monte Carlo using the R packages `greta` and `greta.gp` [34**?** ]. Posterior samples of model parameters were generated by 10 independent chains of a Hamiltonian Monte Carlo sampler, each run for 1000 iterations after an initial, discarded, ‘warm-up’ period (1000 iterations per chain) during which the sampler step size and diagonal mass matrix was tuned, and the regions of highest density located. Convergence was assessed by visual assessment of chains, ensuring that the potential scale reduction factor for all parameters had values less than 1.1, and that there were at least 1000 effective samples for each parameter.

Visual posterior predictive checks were performed to ensure that the observed data were consistent with the posterior predictive density over all cases (and survey results), and over time-varying case predictions within each state.

## Data Availability

The analyses performed in the manuscript required access to epidemiological data provided through the Australian National Notifiable Disease Surveillance System (NNDSS), which is not able to be shared under the terms of our data access agreement. Further information on the NNDSS, including how to request access to data from the Australian Government Department of Health, is available at:
https://www1.health.gov.au/internet/main/publishing.nsf/Content/ohp-pub-datasets.htm

## Code Availability

Model code is available at: https://github.com/goldingn/covid19_australia_interventions

## Acknowledgements

Our analyses use surveillance data reported through the Communicable Diseases Network Australia (CDNA) as part of the nationally coordinated response to COVID-19. We thank public health state from incident emergency operations centres in state and territory health departments, and the Australian Government Department of Health, along with state and territory public health laboratories. We thank members of CDNA for their feedback and perspectives on the results of the analyses. This work was directly funded by the Australian Government Department of Health Office of Health Protection. Additional support was provided by the Australian Research Council (NG DECRA fellowship DE180100635) and the National Health and Medical Research Council of Australia through its Centres of Research Excellence (SPECTRUM, GNT1170960) and Investigator Grant Schemes (JMcV Principal Research Fellowship, GNT1117140).

## Ethics statement

The study was undertaken as urgent public health action to support Australia’s COVID-19 pandemic response. The study used data from the Australian National Notifiable Disease Surveillance System (NNDSS) provided to the Australian Government Department of Health under the National Health Security Agreement for the purposes of national communicable disease surveillance. Data from the NNDSS were supplied after de-identification to the investigator team for the purposes of provision of epidemiological advice to government. Contractual obligations established strict data protection protocols agreed between the University of Melbourne and sub-contractors and the Australian Government Department of Health, with oversight and approval for use in supporting Australia’s pandemic response and for publication provided by the data custodians represented by the Communicable Diseases Network of Australia. The ethics of the use of these data for these purposes, including publication, was agreed by the Department of Health with the Communicable Diseases Network of Australia.

## Supplementary Figures and Tables

**Figure S1:**
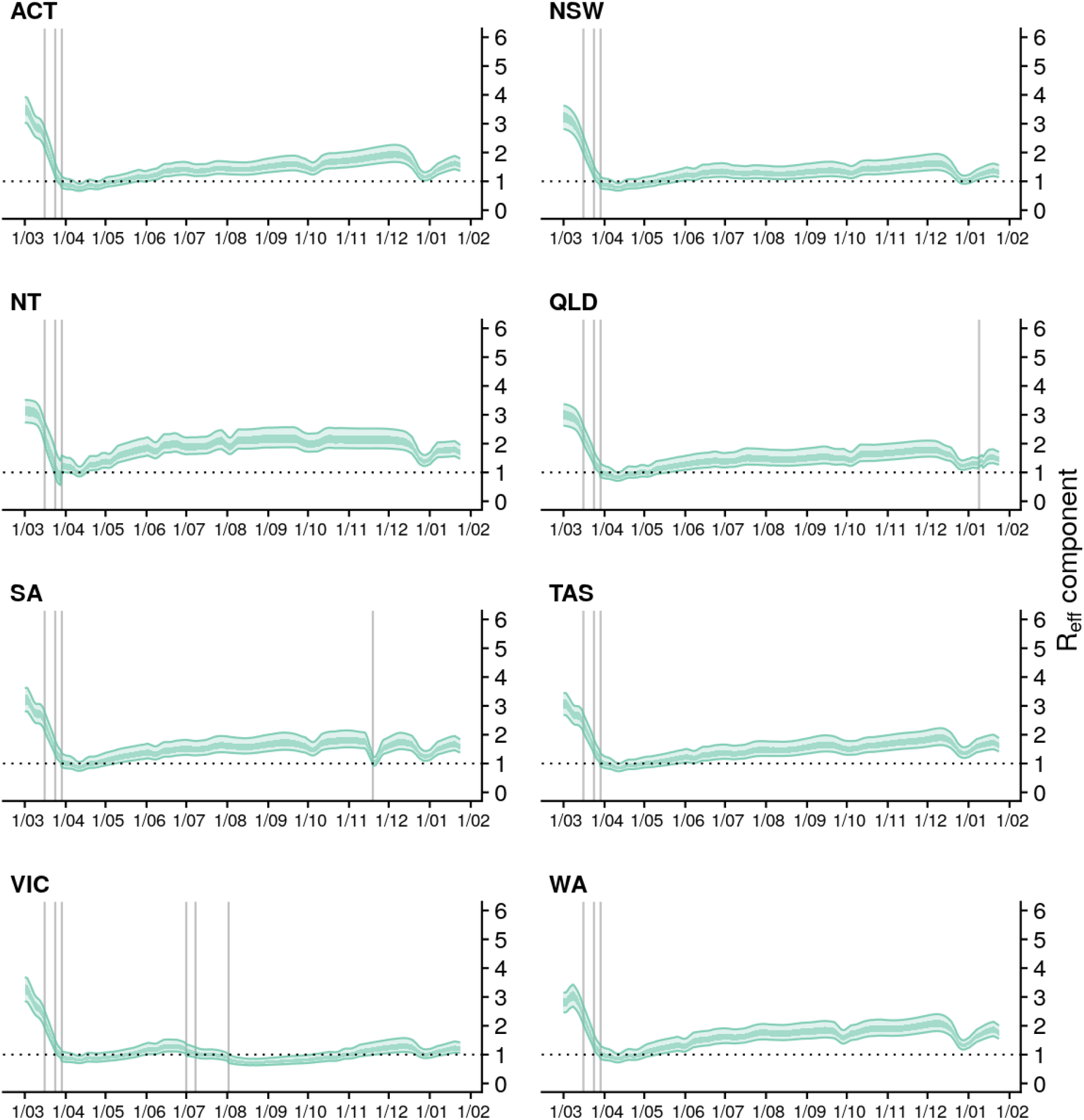
Estimates of state-wide transmission potential (Component 1) by state/territory from 1 March 2020 up to 24 January 2021 (lighter ribbons = 90% credible intervals; darker ribbons = 50% credible interval). Solid grey vertical lines indicate key dates of implementation of various physical distancing policies.

**Figure S2:**
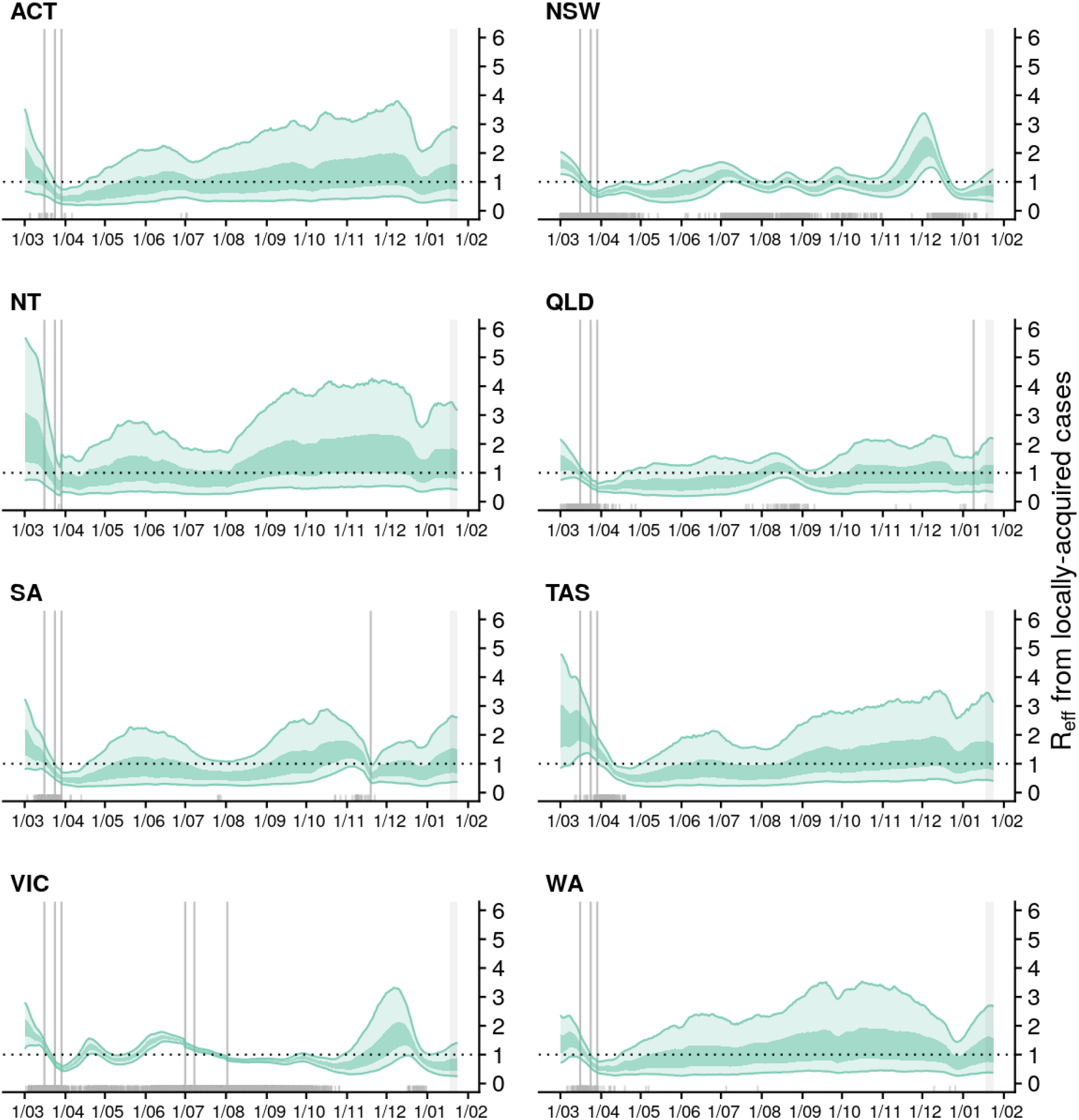
Estimates of *R*_eff_ for local active cases (model Component 1&2) for each state/territory (light green ribbon = 90% credible interval; dark green ribbon = 50% credible interval). Estimates are made from 1 March 2020 up to 24 January 2021 based on cases with inferred infection dates up to and including 18 January (due to a delay from infection to reporting, the trend in estimates after 18 January is informed by our estimates of *R*_eff_ up to 18 January and transmission potential). Solid grey vertical lines indicate key dates of implementation of various physical distancing policies. Black dotted line indicates the target value of 1 for the effective reproduction number required for control. Local cases by inferred date of infection are indicated by grey ticks on the x-axis. For states/territories with very low numbers of local active cases, the estimates of *R*_eff_ for active cases is highly uncertain. The state-wide transmission potential should be referred to when assessing the risk of an epidemic becoming established given a seeding event.

**Figure S3:**
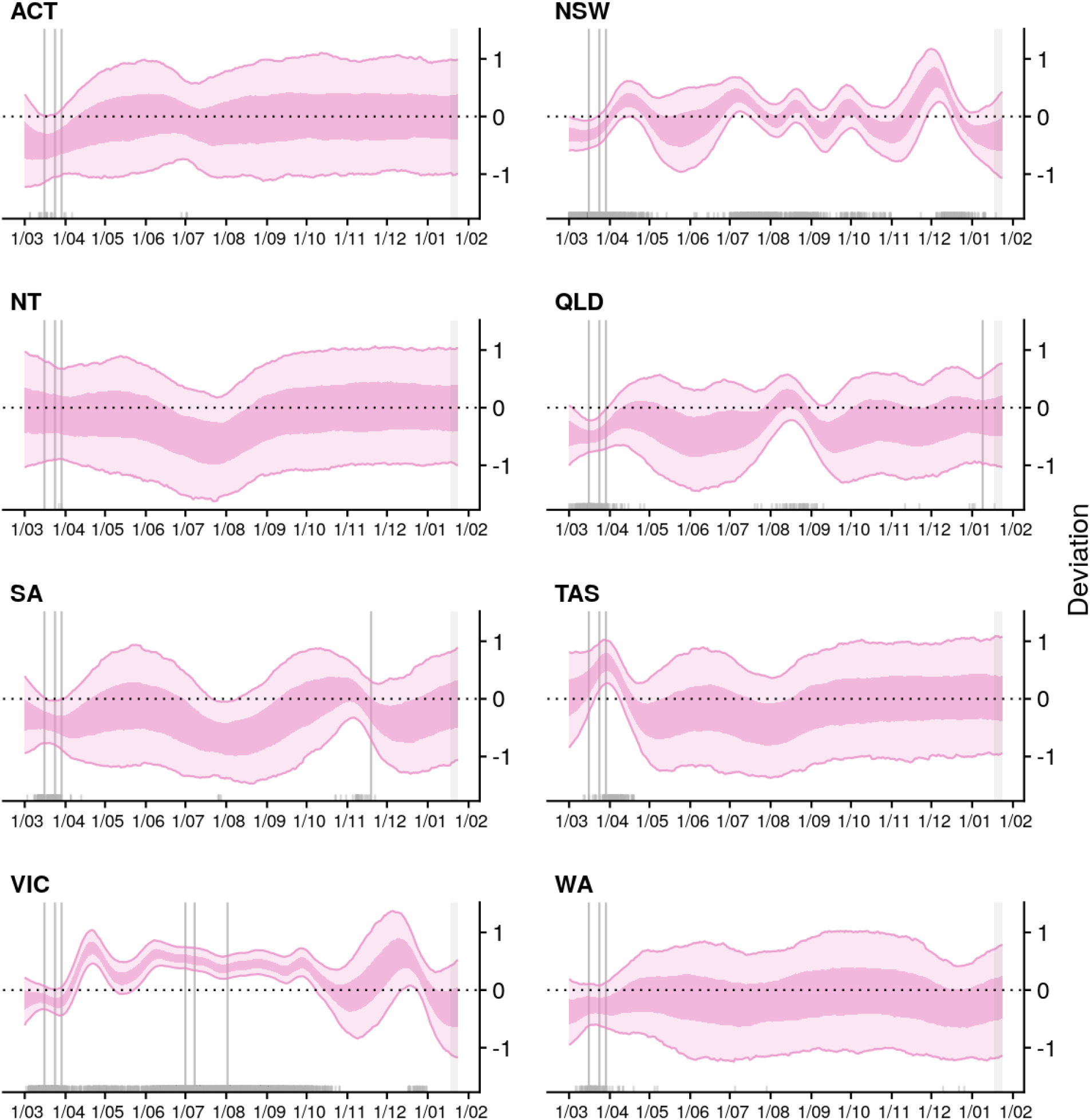
Deviation between *R*_eff_ of active cases and state-level local transmission potential (Component 2) for each state/territory (light pink ribbon = 90% credible interval; dark pink ribbon = 50% credible interval). Estimates are made from 1 March 2020 up to 24 January 2021 based on cases with inferred infection dates up to and including 18 January (due to a delay from infection to reporting, the trend in estimates after 18 January reflects the average range of deviations for that state, indicated by the grey shading). Solid grey vertical lines indicate key dates of implementation of various physical distancing policies. Local cases by inferred date of infection are indicated by grey ticks on the x-axis.

**Figure S4:**
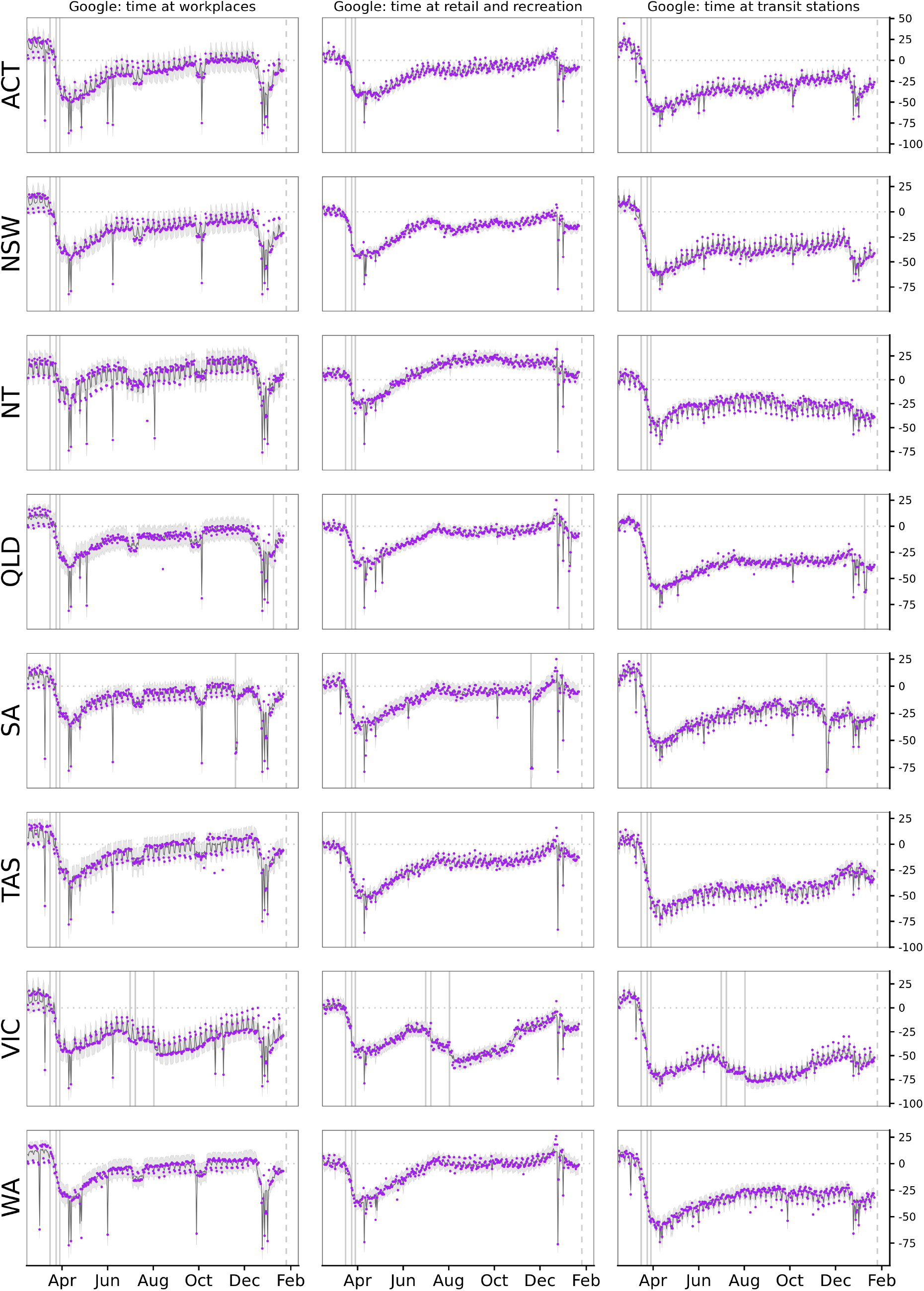
Percentage change compared to a pre-COVID-19 baseline of three key mobility data streams in each Australian state and territory from 1 March up to 24 January 2021. Solid vertical lines indicate dates of implementation of key physical distancing measures. Purple dots in each panel are data stream values (percentage change on baseline). Solid lines and grey shaded regions are the estimated trend and 95% error interval estimated by our model.

**Figure S5:**
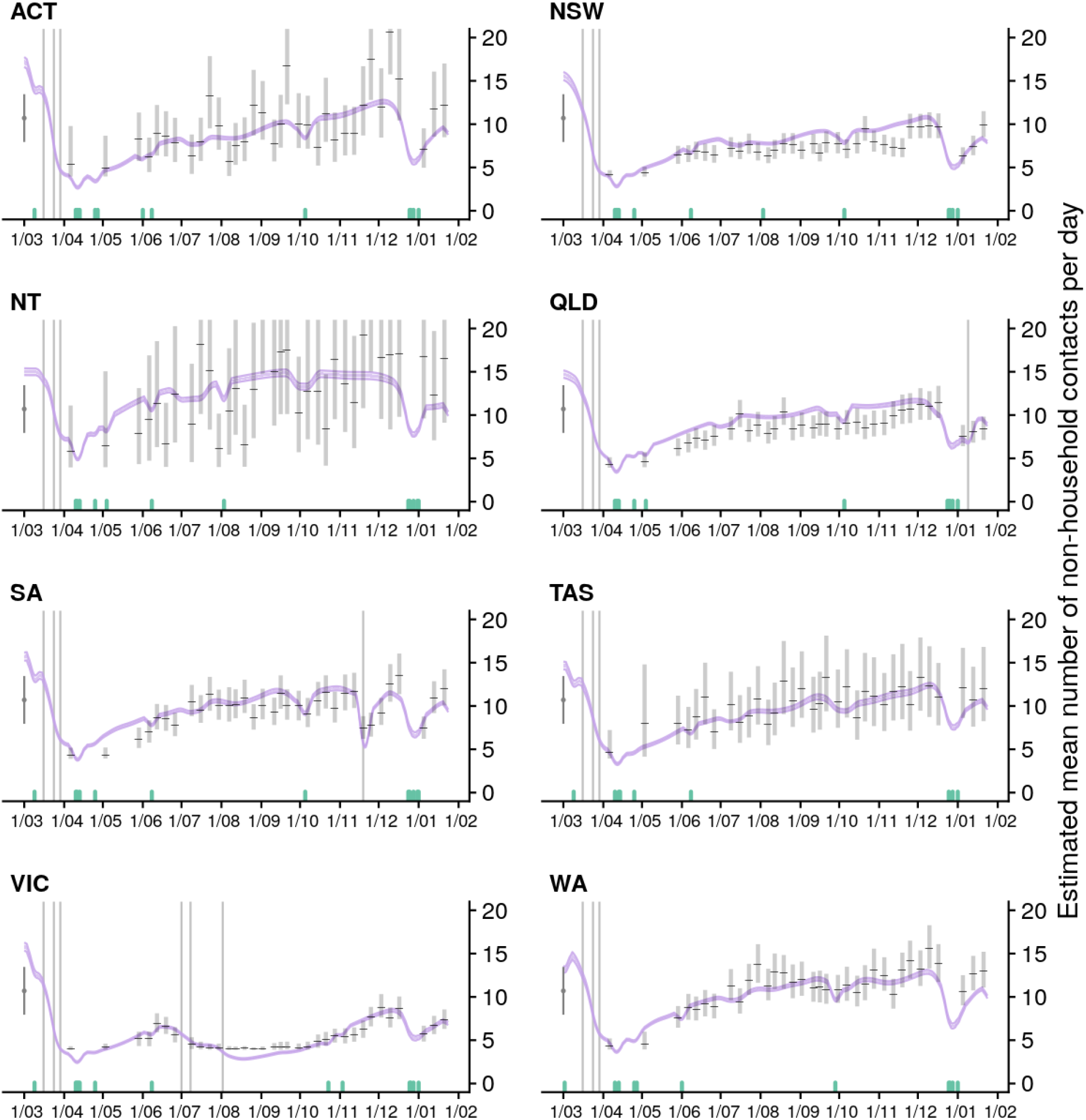
Estimated trend in macro-distancing behaviour, *i*.*e*., reduction in the daily rate of non-household contacts, in each Australian state/territory from 1 March 2020 up to 24 January 2021 (light purple ribbons = 90% credible intervals; dark purple ribbons = 50% credible intervals). Estimates are informed by state-level data from nationwide surveys (indicated by the black lines and grey rectangles) and population mobility data. Green ticks indicate the dates that public holidays coincided with surveys (when people tend to stay home, biasing down the number of non-household contacts reported on those days).

**Figure S6:**
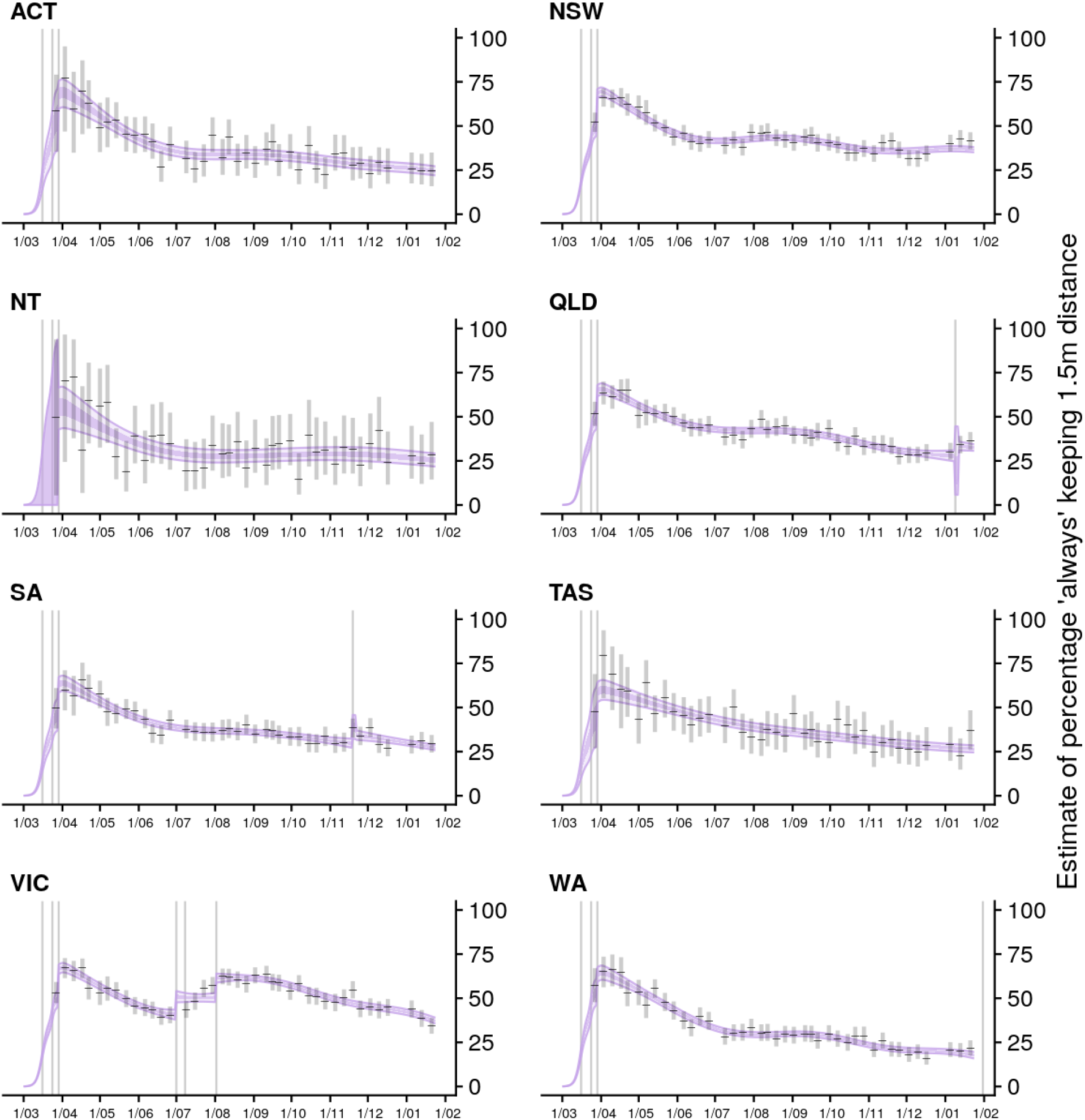
Estimated trend in micro-distancing behaviour, *i*.*e*. reduction in transmission probability per non-household contact, in each Australian state/territory from 1 March 2020 up to 24 January 2021 (light purple ribbons = 90% credible intervals; dark purple ribbons = 50% credible intervals). Estimates are informed by state-level data from nationwide weekly surveys since March 2020 (indicated by the black lines and grey boxes).

**Figure S7:**
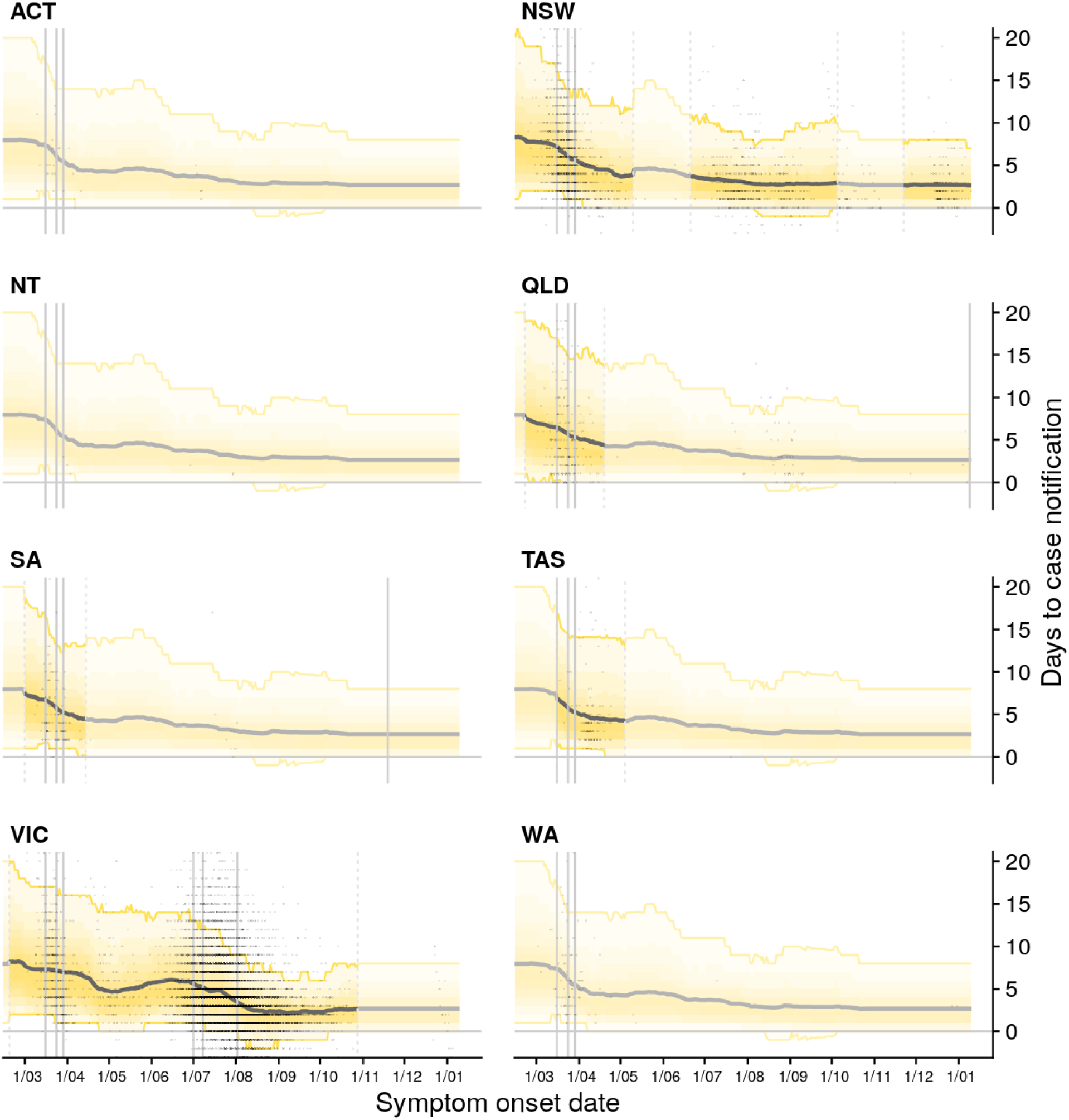
Estimated trend in distributions of time from symptom onset to notification for locally acquired cases for each Australian state/territory from 1 March 2020 to 12 January 2021 (black line = median; yellow ribbons = 90% distribution quantiles; black dots = time-to-notification of each case). Faded regions indicate where a national trend is used due to low case counts.

**Figure S8:**
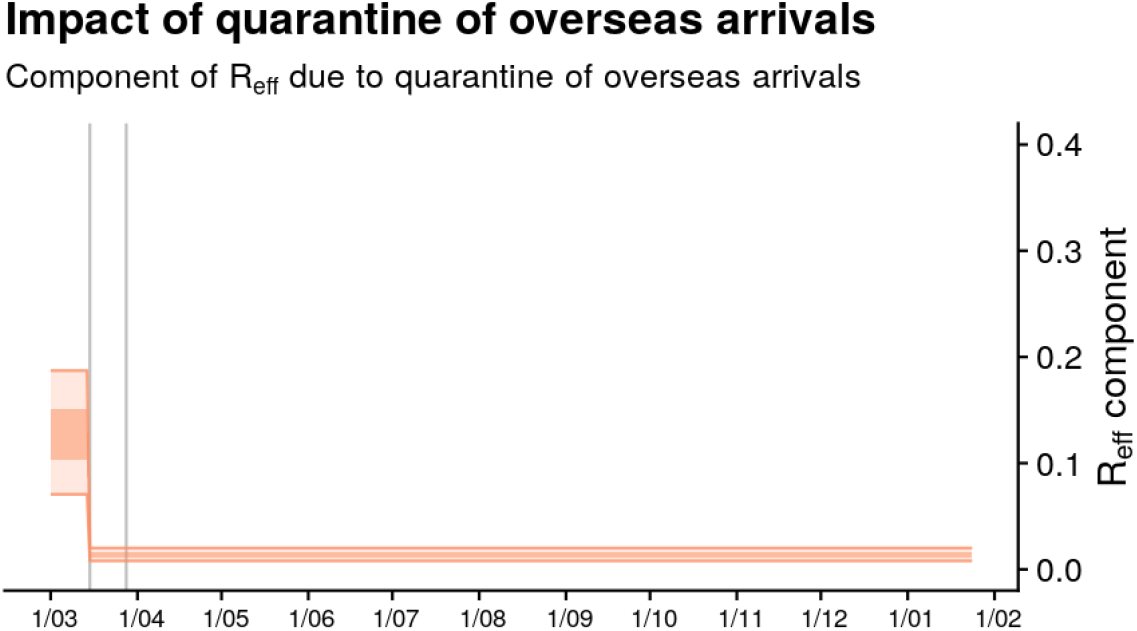
Nationwide average reduction in *R*_eff_ that is due to quarantine of overseas arrivals estimated from the *R*_eff_ model (light orange ribbon=90% credible interval; dark orange ribbon = 50% credible interval). Note that this trend does not capture time-varying fluctuations in *R*_eff_ in each state/territory. Solid grey vertical lines indicate key dates of implementation of key response policies. Black dotted line indicates the target value of 1 for the effective reproduction number required for control. Note: A simple but naïve upper bound on *R*_eff_ import can be computed by assuming that all locally acquired cases arose from imported cases, and therefore computing the ratio of the numbers of local and imported cases. This results in a maximum possible value of the average *R*_eff_ import of 0.57.

**Table S1:** Dates of key changes in restrictions on gatherings and movement for New South Wales and Victoria, as shown in Figures 2 and 3.

| State | Measure | Date |
| --- | --- | --- |
| NSW | Restriction of gatherings to 500 people or fewer | 2020-03-16 |
|  | Closure of pubs, cafes and restaurants | 2020-03-23 |
|  | Stage 3 stay-at-home restrictions imposed | 2020-03-31 |
|  | e. Lifting of stay-at-home restrictions, up to 5 people can gather in households and 10 outdoors, and cafés and restaurants re-open (up to 10 customers) | 2020-05-15 |
|  | Restrictions tightening on pubs | 2020-07-17 |
|  | Restrictions on cafes, restaurants, funerals and weddings | 2020-07-24 |
|  | Restrictions eased on recreation and entertainment facilities | 2020-08-28 |
|  | Stay-at-home restrictions for selected postcodes | 2020-12-19 |
|  | Lifting of stay-at-home restrictions for selected postcodes | 2021-01-09 |
| VIC | Restriction of gatherings to 500 people or fewer | 2020-03-16 |
|  | Closure of pubs, cafes and restaurants | 2020-03-23 |
|  | Stage 3 stay-at-home restrictions imposed | 2020-03-30 |
|  | Lifting of stay-at-home restrictions, up to 5 people can gather in households and 10 outdoors | 2020-05-13 |
|  | Restrictions tightened on household (up to 5 people) and outdoor (up to 10 people) gatherings | 2020-06-22 |
|  | Stay-at-home restrictions imposed in selected Melbourne postcodes | 2020-07-02 |
|  | Stage 3 stay-at-home restrictions imposed in metropolitan Melbourne | 2020-07-09 |
|  | Stage 4 stay-at-home restrictions imposed in metropolitan Melbourne including curfew and travel radius of 5km | 2020-08-02 |
|  | Lifting of stay-at-home restrictions, cafes, restaurants and pubs re-open | 2020-10-28 |

**Table S2:** Parameters in the semi-mechanistic part of the time-varying model of *R*_eff_. Prior on weights for *ω* correspond to Google mobility metrics in the following order: parks, residential, retail and recreation, transit stations, workplaces.

| Prior distribution | Parameter description |
| --- | --- |
| $r^{-1/2} \sim N^+(0, 0.5)$ | Overdispersion of observed daily new infections |
| $\text{logit}(p) \sim N(2.57, 0.08^2)$ | Transmission probability per hour contact time |
| $HC_0 \sim N^+(2.09, 0.06^2)$ | Baseline average daily household contacts |
| $NC_0 \sim N^+(10.70, 0.28^2)$ | Baseline average daily non-household contacts |
| $HD_0 \sim N^+(1.05, 1.68^2)$ | Baseline daily duration per household contact (hours) |
| $ND_0 \sim N^+(0.687, 0.05^2)$ | Baseline daily duration per non-household contact (hours) |
| $\omega \sim N^+(0, 1^2)$ | Mobility-metric weights for non-household contact rates |
| $\alpha \sim N(0, 1)$ | Effect of day of the week on non-household contact rates |
| $\theta \sim N(0, 1)$ | Effect of day-of-week/mobility interaction on contact rate responses |
| $r_{NC}^{-1/2} \sim N^+(0, 0.5)$ | Overdispersion of daily non-household contacts |
| $\mu_{\kappa_{i,0}} \sim N(0, 10^2)$ | Hierarchical mean for peak microdistancing timing |
| $\mu_{\kappa_{i,l}} \sim N(0, 10^2)$ | Hierarchical mean for microdistancing inflection timing |
| $\mu_{h_{i,l}} \sim N(0, 10^2)$ | Hierarchical mean for microdistancing inflection height |
| $\sigma_{\kappa_{i,0}} \sim N^+(0, 0.5^2)$ | Hierarchical s.d. for peak microdistancing timing |
| $\sigma_{\kappa_{i,l}} \sim N^+(0, 0.5^2)$ | Hierarchical s.d. for microdistancing inflection timing |
| $\sigma_{h_{i,l}} \sim N^+(0, 0.5^2)$ | Hierarchical s.d. for microdistancing inflection height |
| $\beta \sim U(0, 1)$ | Microdistancing effect on transmission |
| $q_1 \sim U(0, 1)$ | Effect of quarantine of overseas arrivals (phase 1) |
| $q_2 \times q_1 \sim U(0, 1)$ | Relative effect of quarantine (phase 2 vs 1) |
| $q_3 \times q_2 \sim U(0, 1)$ | Relative effect of quarantine (phase 3 vs 2) |

**Table S3:**
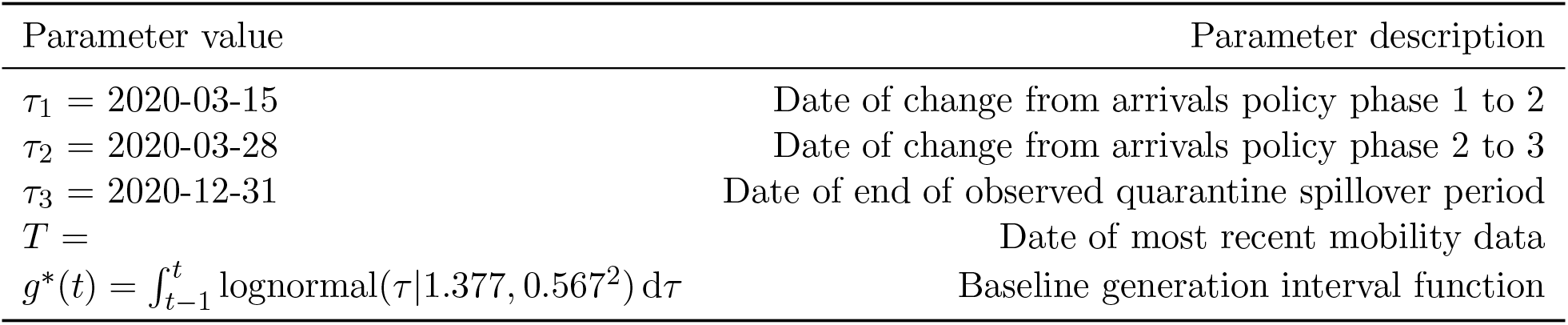
Fixed parameters in the semi-mechanistic part of the time-varying model of *R*_eff_.

**Table S4:** Parameters used in the time-series part of the time-varying model of *R*_eff_.

| Prior distribution | Parameter description |
| --- | --- |
| $\sigma \sim N^+(0, 0.5^2)$ | State-level component of amplitude of deviation $R_{\text{eff}}$ |
| $l \sim \text{lognormal}(3, 1)$ | Temporal correlation $R_{\text{eff}}$ |
| $\alpha \sim \text{lognormal}(3, 1)$ | Correlation mixture weights $R_{\text{eff}}$ |

## Notes

### Competing Interest Statement

The authors of this manuscript are contracted by the Australian Government Department of Health to provide epidemiological analysis to support the COVID-19 response. James McCaw and Jodie McVernon are invited expert members of the Australian Health Protection Principal Committee.

### Author Declarations

The study was undertaken as urgent public health action to support Australia's COVID-19 pandemic response. The study used data from the Australian National Notifiable Disease Surveillance System (NNDSS) provided to the Australian Government Department of Health under the National Health Security Agreement for the purposes of national communicable disease surveillance. Data from the NNDSS were supplied after de-identification to the investigator team for the purposes of provision of epidemiological advice to government. Contractual obligations established strict data protection protocols agreed between the University of Melbourne and sub-contractors and the Australian Government Department of Health, with oversight and approval for use in supporting Australia's pandemic response and for publication provided by the data custodians represented by the Communicable Diseases Network of Australia. The ethics of the use of these data for these purposes, including publication, was agreed by the Department of Health with the Communicable Diseases Network of Australia.

